# Complete genomic profiles of 1,496 Taiwanese reveal curated medical insights

**DOI:** 10.1101/2021.12.23.21268291

**Authors:** Dung-Chi Wu, Jacob Shu-Jui Hsu, Chien-Yu Chen, Shang-Hung Shih, Jen-Feng Liu, Ya-Chen Tsai, Tung-Lin Lee, Wei-An Chen, Yi-Hsuan Tseng, Yi-Chung Lo, Hong-Ye Lin, Yi-Chieh Chen, Jing-Yi Chen, Darby Tien-Hao Chang, Wei-Hong Guo, Hsin-Hsiang Mao, Pei-Lung Chen

## Abstract

**Background:** Taiwan Biobank (TWB) project has built a nationwide database to facilitate the basic and clinical collaboration within the island and internationally, which is one of the valuable public datasets of the East Asian population. This study provided comprehensive genomic medicine findings from 1,496 WGS data from TWB.

**Methods:** We reanalyzed 1,496 Illumina-based whole genome sequences (WGS) of Taiwanese participants with at least 30X depth of coverage by Sentieon DNAscope, a precisionFDA challenge winner method. All single nucleotide variants (SNV) and small insertions/deletions (Indel) have been jointly called and recalibrated as one cohort dataset. Multiple practicing clinicians have reviewed clinically significant variants.

**Results:** We found that each Taiwanese has 6,870.7 globally novel variants and classified all genomic positions according to the recalibrated sequence qualities. The variant quality score helps distinguish actual genetic variants among the technical false-positive variants, making the accurate variant minor allele frequency (MAF). All variant annotation information can be browsed at **TaiwanGenomes** (https://genomes.tw). We detected 54 PharmGKB-reported Cytochrome P450 (*CYP*) genes haplotype-drug pairs with MAF over 10% in the TWB cohort and 39.8% (439/1103) Taiwanese harbored at least one PharmGKB-reported human leukocyte antigen (*HLA*) risk allele. We also identified 23 variants located at ACMG secondary finding V3 gene list from 25 participants, indicating 1.67% of the population is harboring at least one medical actionable variant. For carrier status of all known pathogenic variants, we estimated one in 22 couples (4.52%) would be under the risk of having offspring with at least one pathogenic variant, which is in line with Japanese (JPN) and Singaporean (SGN) populations. We also detected 6.88% and 2.02% of carrier rates for alpha thalassemia and spinal muscular atrophy (SMA) for copy number pathogenic variants, respectively.

**Conclusion:** As WGS has become affordable for everyone, a person only needs to test once for a lifetime; comprehensive WGS data reanalysis of the genomic profile will have a significant clinical impact. Our study highlights the overall picture of a complete genomic profile with medical information for a population and individuals.

## Background

As the concepts of precision medicine prevail, many population-based consortiums have been collecting genomic information for biomedical studies [1]. In Asia, a genetic reference panel based on 3,552 individuals of the Japanese (JPN) population was published in 2019 [2] [3].

Another study based on 4,810 Singaporeans (SGN) was published in 2019 by the SG10K project [4]. The results showed that the population diversity could be captured from a significant sample size cohort analysis. They discovered more than 50 million novel variants. Even though the ChinaMAP project discloses the genetic profile of 10,588 Chinese [5], the Asian genomics data are still relatively underrepresented in the public databases considering the population size worldwide. The Taiwan Biobank project (TWB) has been established to facilitate biomedical research on the genetic basis of Taiwanese, a multicultural population. The majority of Taiwanese immigrated from many provinces of China over the past hundreds of years. Besides, there is a group of Taiwanese aboriginals. As of 30 Sep 2021, TWB has already recruited 153,543 subjects from the general population. Many types of genomic data are available upon application, including single nucleotide variant (SNV) array data from 114,604 subjects, whole-genome sequencing (WGS) data from 2,010 subjects, and the human leukocyte antigen (HLA) typing data from 1,102 subjects. Even though many genotyping approaches can reveal specific variants with disease susceptibility [6], fragmented variant information is not sufficient to fully infer the haplotype of a gene. Whole-genome sequencing data has the potential to reconstruct haplotype information in high resolution, and WGS data covers entire genomic regions allowing complete ascertainment of disease-causing variants, including SNV, small insertion/deletion, structure variants, and the haplotype of a gene. For example, a previous study suggested that more than 95% of hemoglobin subunit alpha (*HBA1/2)* pathogenic or likely pathogenic variants are from *–SEA*, *-a3.7*, *-a4.2* in the Chinese population [7]. Most of them are large deletions with more than 3kb in size. The pathogenic variants of fragile X-linked mental retardation gene FMRP translational regulator 1 (*FMR1)* are the types of trinucleotide (CGG) repeat expansion, for which a specific algorithm is needed to detect the variations in sequencing data [8] [9] [10]. A medical meaningful complete genomic profile of an individual should also include the monogenic disease-causing allele screening [11] and the haplotype of the human leukocyte antigen (*HLA*) gene. Like the *HLA* gene, Cytochrome P450 2D6 (*CYP2D6*) gene is associated with many drug metabolism. For functional interpretation, complete haplotype information is needed for both *HLA* and *CPY2D6* genes.

Identifying at-risk individuals with medically actionable information from hereditary diseases could benefit the whole family. Thus, expanded carrier screening for monogenic diseases has become a common consensus since a joint statement published by The American College of Medical Genetics and Genomics (ACMG), The American College of Obstetricians and Gynecologists (ACOG), The National Society of Genetic Counselors (NSGC), Perinatal Quality Foundation (PQF), and Society for Maternal-Fetal Medicine (SMFM) in 2015 [12].

All the listed disorders involve a cognitive or physical disability, the need for postnatal surgical or medical intervention, or a detrimental effect on the quality of life; and most importantly, they are disorders where prenatal intervention could significantly improve perinatal outcomes and delivery management or where prenatal education could meet the unique needs after birth. Each year ACMG Secondary Findings Maintenance Working Group (SFWG) evaluates these actionable genes and associated conditions by their actionability, severity, penetrance, and impact or burden of available treatment modalities or screening recommendations. The latest version, ACMG SF v3.0 [13] published in May 2021. It contains 73 actionable genes, in contrast to 59 genes in the previous version. Many studies disclosed the profile of actionable genes in the Asia-Pacific region [14–16]; WGS data allowed us to reanalyze when the gene list has been updated. The advent of WGS helps characterize population genetic structure that can reveal evidence to the public healthcare system, mitigating the cost through disease risk prediction, diagnosis, and treatments. In this study, we reanalyzed the WGS data from 1,496 participants of the TWB to provide medically helpful information on the population’s complete genomic profile. We also discussed disease carrier status, including all the conditions curated in the ClinVar database, ACMG actionable genes, drug responses from the PharmGKB database, and the clinically relevant variant with high MAF in the Taiwanese population. To facilitate exploration of the variants disclosed in this study, we designed a web interface for the constructed database TaiwanGenomes (https://genomes.tw) for easily browsing the list of variants under different combinations of filters.

## Methods

### Data source

All 1,496 Taiwanese WGS data were collected and de-identified at Taiwan Biobank (TWB). WGS were conducted separately in three batches (n = 496, 497, and 503, respectively) with high-depth mapped reads. The samples were sequenced by Illumina HiSeq 2500, 4000, and Novaseq system. Sequencing of DNA extracted from each blood sample generated about 90 GB of data with an average coverage of 30x.

### Genotype calling, validation, and variant annotation

Variant detection and joint genotype calling analyses were conducted based on the Sentieon DNAscope pipeline (Sentieon Inc., version 201808 [17]), which is a commercial implementation of GATK’s best practice [18]. The sequence reads in FASTQ format of each sample were aligned against the human reference genome (GRCh37/ucsc.hg19.fasta) using BWA-MEM (Burrows-Wheeler Aligner with Maximal Exact Match algorithm, version 0.7.15-r1140 [19]). The alignment file was sorted by Samtools [20], and the Sentieon Dedup marked the duplicated reads. The Sentieon Realigner reinforced local realignment around each indel region, and the Sentieon QualCal recalibrated base quality scores. Single nucleotide variants (SNV) and short insertions/deletions (INDEL) were called in genomic variant call format (GVCF) by Haplotyper. The Sentieon GVCFtyper jointly called 1,496 subjects as a cohort, then Sentieon VarCal plus ApplyVarCal, a machine-learning-based variant quality sequence recalibration (VQSR) algorithm for variant refinement. The training sets for VQSR were identical to the GATK’s best practice. For SNV, the datasets from the 1000G phase1, omni2.5, dbSNP version 138, and HapMap version 3.3 were included as the training sets. For INDEL, the datasets from the 1000G phase1, dbSNP version 138, and Mills-and-1000G gold standard were used as the training sets. VQSR included a list of sequence level annotations such as QD, MQ, MQRankSum, ReadPosRankSum, and FS. Multi-allelic variants were normalized into multiple bi-allelic variants at the same position through decomposition and left-aligned using bcftools (v1.9) [20, 21]. We repeated the same pipeline with an additional seven Genome in A Bottle (GIAB) samples (HG001∼HG007) and jointly called with 1,496 TWB subjects for benchmarking the variant calling pipeline. We stratified the variant call sets according to the VQSR tranche 100.0, 99.9, 99.8, 99.7, 99.6, 99.5, and 99.0. We further adopted the call rate, allele number (AN), and depth of coverage (DP) criteria for classifying all reference alleles into three quality classifications. Moreover, we randomly validated hundreds of variants in four samples (NGS2_20150510B, NGS2_20150510C, NGS2_20150510D, NGS2_20150510F) by Sanger sequencing. We are particularly interested in the variants absent after joint calling or only present after joint calling. Due to limited DNA, we separated samples into two groups (BC and DF) for cross-validation. We utilized the Sanger sequencing results to calculate the estimated false discovery rate (FDR) for evaluating the VQSR tranche. We concluded VQSR tranche 99.7 as PASS by comparing HG001 (NA12878) genetic variants and the results from Sanger sequencing validation. All variants can be accessed from the database **TaiwanGenomes (genomes.tw)** with VQSR tranche and annotated information. Only variants with PASS quality were considered in the downstream analysis.

Variant annotation was done by ANNOVAR (version 20180416) [22] with updated databases, including RefSeq Gene, UCSC Known Gene, ClinVar (v20210501), avsnp150, NHLBI-ESP 6500 exome, 1000 genome, gnomAD genome and exome (v2.1.1), ExAC 65000 exome, Kaviar, cg69, dbnsfp33a, dbscsnv11, gwava, tfbsConsSites, wgRna, targetScanS. We selected the annotated variants on the autosome and sex chromosome as the final variant call-set. We defined the nonsynonymous variants as where a variant’s annotation hits exonic, non-coding RNA (ncRNA) or splicing region. The variation is connected to nonframeshift deletion, frameshift deletion, nonframeshift insertion, frameshift insertion, stopgain, stoploss, or a nonsynonymous single-nucleotide variant (SNV) in exonic variant function.

To further elucidate the possible role of structural variant calling tools, as a trial, we used Manta (version 1.6.0) [23] and AnnotSV (version 3.0.4) [24] to identify and annotate the structural variants (SV) in *HBA1/2* genes (n = 494, a subset of 1,496). Furthermore, the *SMN1/SMN2* copy number was analyzed by SMNCopyNumberCaller (version 1.1.1) [10].

### Variant filtering and classification

We defined the variant absented in any other public database (e.g., without allele frequency or effect annotation) as the “globally novel variant.” The variant that does not exist in any previous samples while sequentially examining the 1,496 samples is the “populational novel variant.” For functional effect analysis, the variants annotated as exonic in refGene were defined as “coding region variants,” whereas the variants annotated as nonsense, splicing, or frameshift in refGene were defined as “loss of function (LOF) variants.” We wrote the in-house scripts to conduct the filtering processes. Public users can also retrieve the same information by setting corresponding filters in TaiwanGenomes (https://genomes.tw).

### Pharmarcogenomics

The *HLA* gene is crucial in precision medicine. It is associated with many adverse drug events, including antithyroid drug-induced agranulocytosis [25]. The clinical variant data in the PharmGKB database (clinicalVariants.tsv, version 20210405) details how genetic variation influences the drug response. It contains a list of variant-drug pairs with the level of evidence. Here, we reported the variant frequency in the TWB cohort for only the variants with the level of evidence 1A, 1B, and 2A variants. There are 1,103 subjects with detailed *HLA* types (https://taiwanview.twbiobank.org.tw/hla) by TWB, in which 884 subjects are the subset of 1,496. We did not include the remaining 219 subjects whose WGS were sequenced by the ThermoFisher Ion Proton platform. To evaluate the allele frequencies of PharmGKB-reported *HLA* variant-drug pairs in the Taiwan population, we queried *HLA* typing from the TWB websites. For *CYP* genes, we genotyped *CYP* haplotypes based on 1,017 WGS data by ALDY [26] and STARGAZER [27] (a subset of 1,496, manuscript in preparation). In calculating *CYP* variant-drug pairs, we only reported the allele frequency of the variants associated with abnormal drug metabolism. We did not include *CYP* wild-type allele frequencies.

### Carrier status – cohort MAF and expert reviews

ClinVar [28] database collects the interpretation of clinically related variants submitted by global researchers. Many known monogenic disease-causing variants are rare and vary across populations [29]. However, only a few records in the database are from the Asian population. To explore the medical relevant genetic variants in the Taiwanese people, we retained the annotated variants as pathogenic, likely_pathogenic, or pathogenic/likely_pathogenic in ClinVar (v20210501). A high variant minor frequency (MAF) allele ( > 0.5%) with known pathogenicity might indicate the specificity of the population’s genetic architecture. By comparing across the East Asian population, we singled out the pathogenic or likely-pathogenic variants with high-frequency try to address the specific genetic structure of Taiwanese.

The variant MAF was also considered by comparing their allelic frequencies in the gnomAD database (version 2.1.1) and the ExAC database. The following filtration criteria were used:

1. The minor allele frequency of the variant in TWB1496 was ≥ 0.01, while that in the ExAC database and the East Asia population of gnomAD genome and exome database were ≤ 0.01 or not seen, 2) the allelic frequency is between 0.005 and 0.01 in TWB1496, while that in the ExAC database and the East Asia population of gnomeAD genome and exome database were ≤ 0.01 or not seen. The genetic inheritance mode of the variants was manually annotated using the Online Mendelian Inheritance in Man database (OMIM). Experts further reviewed the variants within ACMG actionable genes. The concept of medically actionable genes originated from ACMG recommendations on reporting secondary findings from exome or genome sequencing results. To further focus on the East Asian population, we also compared the allele frequency of variants in the carrier sets to two East Asian populations, e.g., Singaporean and Japanese. The Singaporean data was acquired from the SG10K project [4], and the Japanese data was obtained from the 3.5KJPN [30].

To evaluate the clinical impact of the carrier call set, we applied the expanded carrier panel [31] to the above data to calculate accumulation frequencies of pathogenic and likely pathogenic variants in monogenic disease-causing genes. We compared these data with the United States cohort[31](sequencing method) and Taiwan domestic data [6] (array genotyping method). The overall study design and analysis workflow are shown in Figure S1.

We also used the same carrier call-set data to evaluate other monogenic disease-causing genes as a concept of expanded carrier screening. We compared the pathogenic/likely pathogenic variant carrier frequency with the U.S. cohort [31] and Taiwanese cohort [6] (using TWBv2.0 array genotyping). Moreover, we also used the 274-gene list [31] as a virtual panel to estimate how many couples may benefit from the expanded carrier screening in the Taiwan population.

### Web application for TWB1496 cohort

The web application for browsing **TaiwanGenomes** is based on an open-source project—VASH (https://github.com/mbilab/vash). VASH, a composite word of variant and flash, aims to provide a rapid and fluent user experience while browsing whole genome-scale variants. VASH is implemented using Vue.js (https://vuejs.org/) and Django (https://djangoproject.as the frontend frameworks, respectively. MySQL (https://www.mysql.com/) is used as the database engine of Django. All the requests of VASH are segmented into small chunks and well cached through the entire processing pipeline, from Vue.js to MySQL. Therefore, VASH is capable of processing the following few chunks while users are viewing previous chunks. VASH enables infinite scrolling, which is more intuitive than pagination browsing and achieves second-response time regardless of the number of queried variants.

## Results

### High-quality variant call sets

To evaluate the accuracy of the joint calling, we used the same pipeline to analyze seven samples (HG001∼HG007) from Genome in A Bottle (GIAB) as the internal quality control. We jointly called 1,496 TWB subjects with seven reference samples and stratified the joint call set according to the VQSR tranche 100.0, 99.9, 99.8, 99.7, 99.6, 99.5, and 99.0. After that, we confirmed both SNV and INDEL variant calling accuracy by comparing HG001 (NA12878) genetic variants as the ground truth to the subset of HG001 results in the joint called cohort for every variant. We defined VQSR tranche 99.7 as PASS criteria to balance the overall sensitivity and specificity Figure S2(a) and S2(b). All variants can be accessed from the database TaiwanGenomes (genomes.tw) with VQSR tranche and annotated information. We only included variants with PASS quality in all the downstream analyses.

Apart from alternative alleles, we also analyzed reference alleles and used the call rate, allele number (AN), and depth of coverage (DP) filtration for quality classification. We briefly classified all non-alternative positions into A, B, and C categories. If the call rate of a reference allele was above 80% (AN>2,400 & DP>18,000), the reference allele was defined as category A (2,686,586,573 variants). Category B (16,201,674 variants) indicated that the call rate of an allele was below 10% (AN<300 & DP<4,500). Category C (30,369,949 variants) represents the call rates between categories A and B. The researchers should be careful when interpreting the variants in categories B and C as MAF = 0 in the Taiwanese population. The quality information for every genomic locus can be easily browsed at https://genomes.tw/#/supplement.

We selected 18,624 high-quality variants in TWB1496 and gnomAD East Asian for regression analysis. To present the allele frequencies distribution between TWB1496 and gnomAD East Asian, we used the Python package Scipy and Statsmodels.api to compute the linear regressions and scatter plots. We demonstrated the correlation of MAF among different VQSR tranches, respectively (Figure S3A, S3B, S3C, S3D). The r-squared implicated the higher VQSR specificity, the higher correlation to the current gnomAD East Asian dataset.

Variant detection bias or population structure could affect the MAF differences between the two cohorts.

### Sanger confirmation

The differences between the jointly-called VCF file and individual variant calls are worth noting. Theoretically, VQSR was designed to rescue variants with moderate individual-level quality and improve the overall calling accuracy when the sample size increases. Hundreds of variants have been checked by the Sanger sequencing, but only 109 variants matching the following criteria were included for the accuracy analysis: 1) unambiguous biallelic variants; clear Sanger results; 3) inconsistent results between joint-called and individual-called.

Notably, the number of true negative variant calls (TN) was underestimated because the VCF file only included the positive variants. We defined VQSR tranche 99.7 as PASS, which was more accurate than the variant calls with a higher VQSR tranche. By setting VQSR 99.7 as PASS, the false discovery rate (FDR) was reduced by 25% (0.75 to 0.5). Based on the sanger validation analysis, applying VQSR can remove false positive (FP) variants by 68.6% (35/51) (Table S1)

**Table 1.** Variant statistics of the 1,496 WGS from Taiwan Biobank

|  |  | 1,496 WGS |  |  |
| --- | --- | --- | --- | --- |
|  |  | ALL | PASS | PASS / Globally novel |
| Number of variants after normalization |  | 59,433,212 | 51,135,411 | 16,520,159 |
| Minor allele frequency (MAF) | MAF < 0.05 | 49,701,036 | 42,557,774 | 16,066,996 |
|  | MAF < 0.01 | 44,591,573 | 38,599,450 | 15,495,346 |
| Coding regions | All | 480,784 | 439,192 | 26,664 |
|  | Nonsynonymous | 274,265 | 250,940 | 700 |
| Loss of function | Frameshift | 10,225 | 9,528 | 3,174 |
|  | Non-sense | 6,402 | 5,746 | 151 |
|  | Splicing | 6,871 | 5,663 | 834 |

### Novel variants in 1,496 Taiwanese

Our final variant call-set consisted of 59,433,212 variants from autosome and sex chromosome (Table 1). Among these variants, there are 49,701,036 variants with minor allele frequency (MAF) < 0.05 and 44,591,573 variants with MAF < 0.01. There are 480,784 variants in coding regions, and 274,265 variants are annotated as non-synonymous. For the PASS variants with the criterion VQSR 99.7, the number of variants decreased to 51,135,411. Among them, there are 42,557,774 variants with MAF < 0.05 and 38,599,450 variants with MAF < 0.01. In addition, there are 439,192 variants in coding regions and 250,940 variants are annotated as non-synonymous.

To analyze the unique genetic characteristic of the Taiwanese population, we further filtered out the variants that did not exist in other databases (see methods). There are 16,520,159 variants classified as “globally novel variants.” To investigate the population genetic structure further, we sorted the sample by their total variant numbers and calculated the unique globally novel variants (Fig. 1). The results showed that a Taiwanese has 6,870.7 globally novel variants on average. Overall, there are 16,066,996 variants with MAF < 0.05 and 15,495,346 variants with MAF < 0.01. Most of these novel variants are in the intergenic regions. There are 26,664 variants in coding regions and 700 annotated as missense variants. Moreover, 4,159 were annotated as loss-of-function variants, including 151 nonsense variants, 3,174 frameshift variants, and 834 splicing variants.

**Figure 1.**
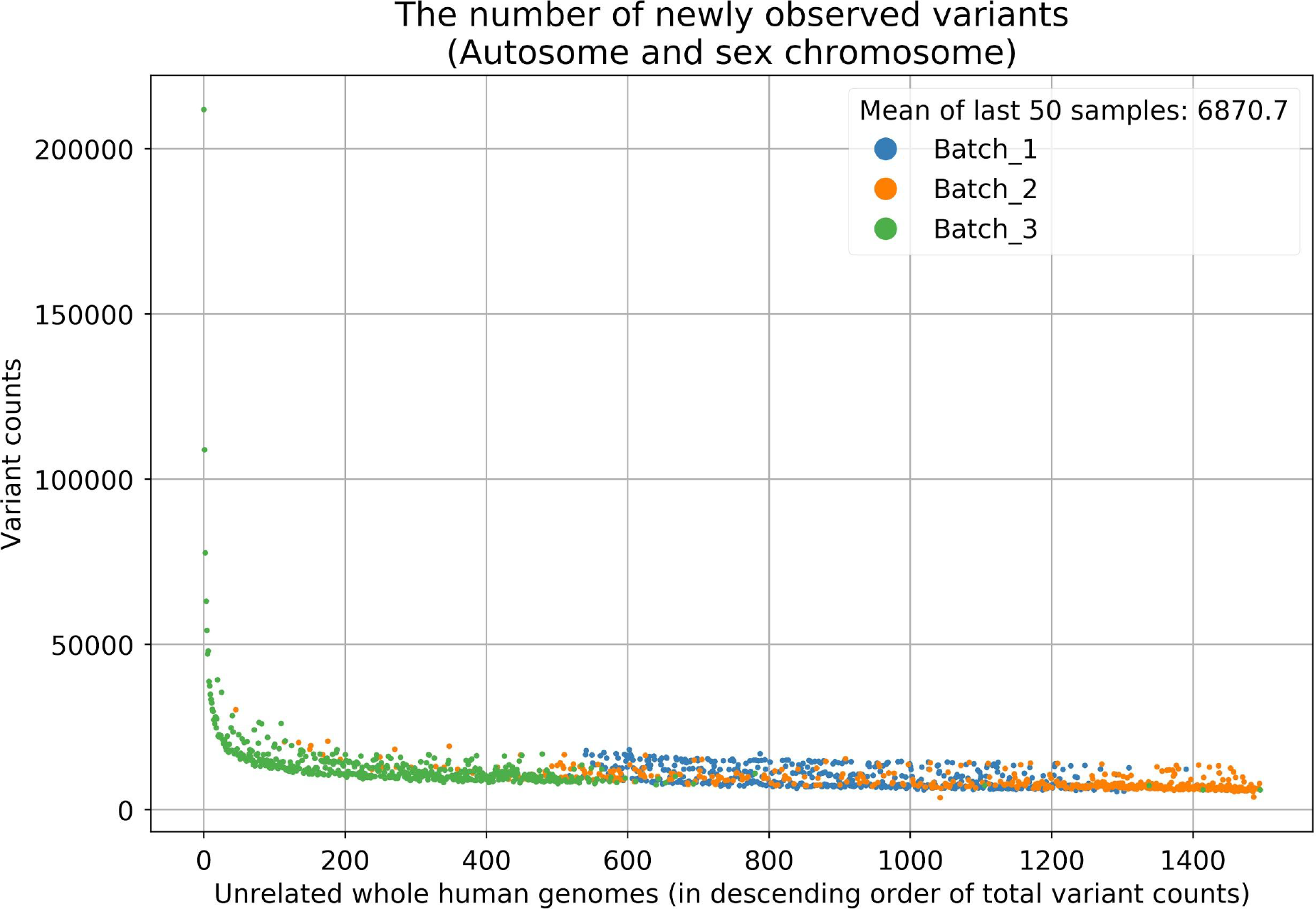
Novel variants per individual. Novel variants per individual. As more individuals are sequenced, the number of newly observed variants in an individual dramatically decreases. The mean of the populationally novel variants among the last 50 individuals is 12,364.10 (VQSR 99.7), and the mean of the globally novel variants among the last 50 individuals is 6,870.70.

Loss-of-function and deleterious variants play a crucial role in Mendelian disorders. From the spectrum of variant numbers and the allele frequencies, we observed that the loss-of-function variants were much fewer than others under negative selection, which is in line with our expectations. We also observed that the occurrences of non-frameshift indels were almost the same as loss-of-function variants. (Fig. 2)

**Figure 2.**
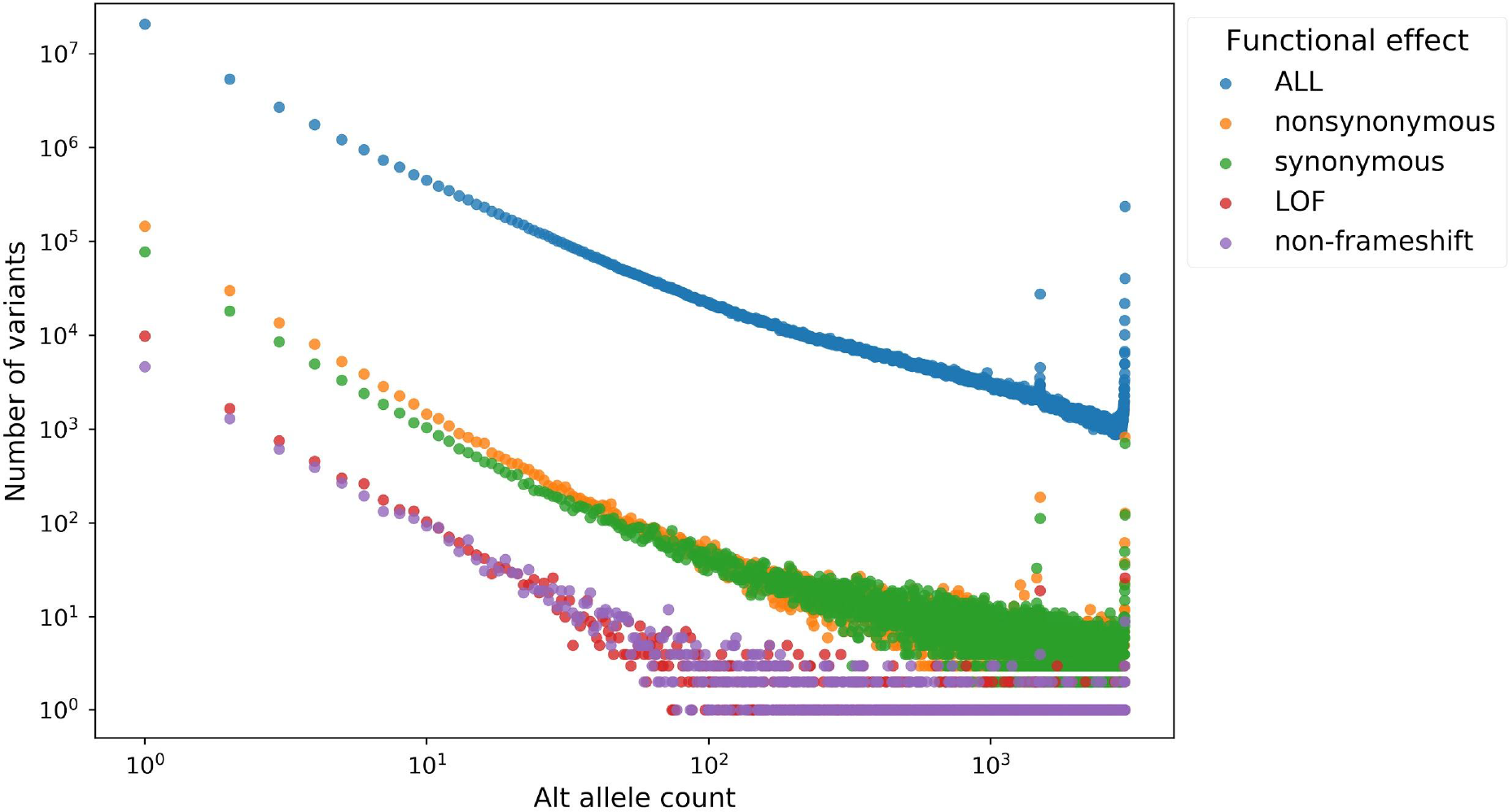
Annotated functional effect and occurrence of all passed variants. The spectrum between the alternative allele counts and the number of variants in different categories. The variants were categorized based on the ANNOVAR ExonicFunc.RefGene annotation. LOF: loss of function, including frameshift_deletion, frameshift_insertion, stopgain, and stoploss. non-frameshift: including non-frameshift_deletion and non-frameshift_insertion. nonsynonymous: non-synonymous_SNV. synonymous: synonymous_SNV.

### Variants within medically actionable genes in 1,496 Taiwanese

We identified 1,136 variants with clinical evidence, termed carrier call-set, and 58 of which were in the 73 ACMG recommended actionable genes [13]. The 58 variants within ACMG actionable genes were further reviewed by experts, resulting in 53 secondary findings. We also filtered the carrier sets of 1,136 variants by the allele frequency. Among the 1,136 variants, there are 78 variants with allele frequency ≥ 0.005. To strengthen and avoid ambiguity of these 78 variants, we filtered out variants that if the allele frequency were also ≥ 0.005 among any East Asian population in public databases, including ExAC_EAS, gnomAD_genome_AF_eas, and gnomAD_exome_AF_eas. The filtered carrier set consists of nine variants. [In TaiwanGenomes, with the filter setting: CLNSIG: Pathogenic, Pathogenic/Likely_pathogenic, Likely_pathogenic, 1136 variants retrieved. +AF>=0.005, 78 variants; +AF>=0.01, 54 variants; ] After review by experts, 53 variants annotated as pathogenic or expected pathogenic are considered secondary findings. Most samples have only one such variant, and the average occurrence is about 5.54% (Table 2). However, the high occurrence mainly arises from three disease genes with autosomal recessive inheritance, including *MUTYH*, *ATP7B*, and *GAA*. The ACMG guideline suggested that two pathogenic or expected pathogenic variants be reported together. None of the samples harbored a second pathogenic variant in TWB 1,496 cohort, indicating the average occurrence is 1.7%. Notably, one pathogenic variant with high occurrence is *PTEN* (NM_000314.6:c.802-2A>T, rs587782455), a splicing-acceptor variant, which did not pass the VQSR threshold. It has been excluded in the carrier of SF v3.0 analysis.

**Table 2.**
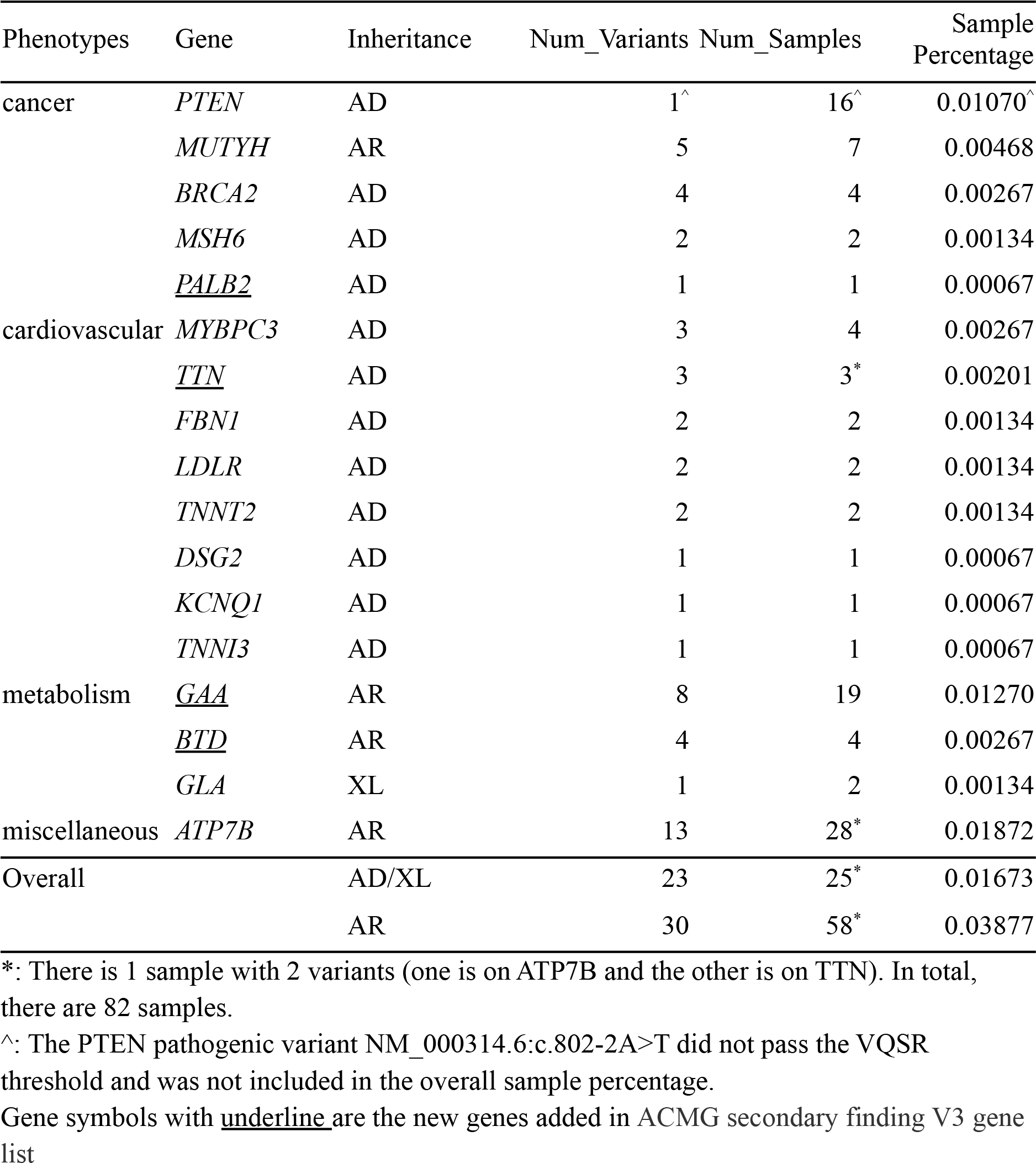
Medically actionable (ACMG SF v3.0) variants in Taiwan Biobank cohort

### Pharmacogenomics loci

For PharmGKB reported clinical variant data with the level of evidence 1A/1B/2A, there were 13 HLA alleles and 17 clinically significant variant-drug pairs. We evaluated the frequencies of 13 HLA haplotypes as risk alleles with matched nomenclature fields from 1,103 TWB subjects (Table S2A). Most HLA haplotypes in PharmGKB are three or four fields, suggesting high-resolution genotypes were necessary. Among 17 variant-drug pairs, we found 16 pairs in the TWB cohort, 13 pairs with haplotype frequencies over 1%. The most common haplotype was HLA-C*01:02:01 group alleles (16.05%), followed by HLA-A*33:03 (12.93%). Two alleles were associated with the adverse drug event (ADE) of taking methazolamide and allopurinol, respectively. However, one known pharmacogenomics allele HLA-DRB*08:03 (8%, 178/2206) was missed in the PharmGKB clinical variant list, suggesting a systematic review is necessary. In 1,103 TWB subjects, we found 439 people carried one risk allele, 147 people had two risk alleles, 170 people carried with three risk alleles, 66 people carried with four risk alleles, and nine people had five risk alleles. HLA haplotype frequency analysis revealed that approximately three out of four Taiwanese people had at least one risk allele, implying a considerable proportion of people can benefit from HLA typing whenever a prescription for a corresponding drug is needed.

In addition, we also evaluated the allele frequencies of PharmGKB-reported CYP variant-drug pairs in the sample of 1,017 TWB volunteers (manuscript in preparation). CYP variants analysis revealed 89 pharmacogenetic variant-drug groups with the level of evidence 1A/1B/2A (Table S2B). Among 89 groups, 54 groups had allele frequencies (excluding WT) ≥ 10% in the TWB cohort. The most common variant was CYP3A5*3 (73.2%), followed by haplotypes of the CYP2C19 group (CYP2C19*2, CYP2C19*3, CYP2C19*9, CYP2C19*10, CYP2C19*17, CYP2C19*24, and CYP2C19*26 )(36.2%). The variants were associated with the abnormal drug metabolism of tacrolimus and omeprazole, implying CYP typing is helpful for clinical medication safety. Treatment alteration occurs when a genetic variant alters treatment’s efficacy, dosage, metabolism, or pharmacokinetics or otherwise causes toxicity or an adverse drug reaction (ADR). This information is precious for both clinicians and patients.

### The status of carrier variants in Taiwan, Singapore, and Japan

To address the uniqueness of Taiwanese, we compared the MAF of 1,136 carrier variants in two different East Asian populations, e.g., Japanese and Singaporean. There were 279 common carrier variants between Japanese and Taiwanese and 611 common carrier variants between Singaporean and Taiwanese (Fig. 3).

**Figure 3.**
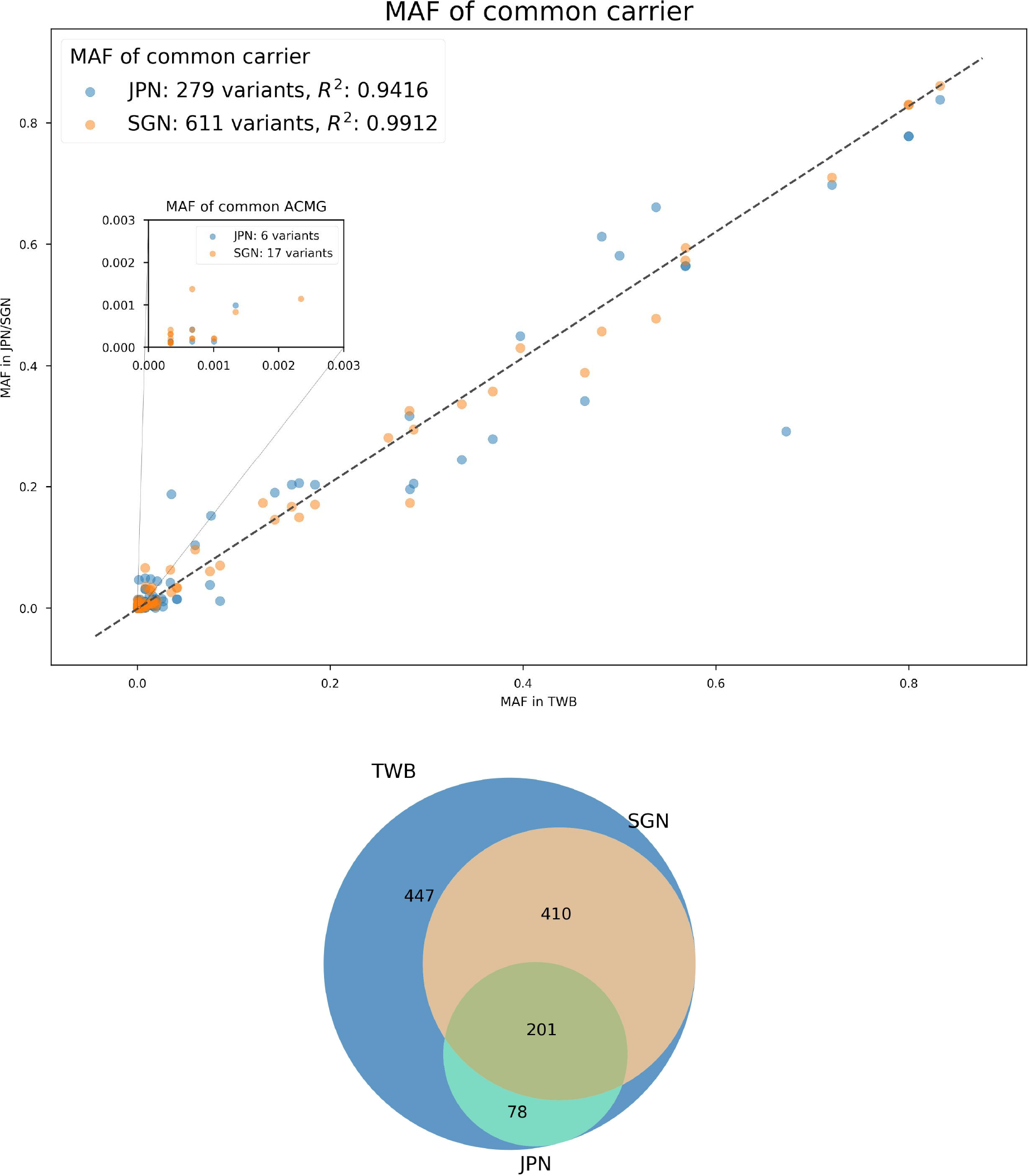
The allele frequencies between TWB, JPN, and SGN populations. X-axis: The minor allele frequency of ClinVar pathogenic/likely pathogenic variants in 1,496 TWB cohort. Y-axis: The minor allele frequency of ClinVar pathogenic/likely pathogenic variants in Japan (JPN) and Singapore (SGN) population. There are 279 carrier variants found both in the TWB and the JPN samples, and there are 611 carrier variants found both in the TWB and the SGN samples. As for 54 acmg secondary findings, there are six variants found in both TWB and JPN, and there are 11 variants found in both TWB and SGN. Venn diagram illustrating the distribution of ClinVar pathogenic/likely pathogenic variants among three populations.

### Carrier status of variants on monogenic disease-causing genes

We compared our carrier data with the United States cohort [31] and another domestic research with the array genotyping method (TWBv2)[6]. The main difference is listed in Table 3A. The most striking difference resides in the *GJB2* gene, a famous hearing impairment contributor. The estimated carrier frequency in the Taiwan biobank NGS database reached 16.7%. More than 90% is contributed from the *GJB2 V37I* variant (8.6%). *GJB2* pathogenic/likely pathogenic (P/VP) carrier frequency in the US cohort is only 6.25% [31], less than one-half of the Taiwan counterpart. In contrast, the *GJB2* carrier rate falls to 1.59% with TWBv2 array genotyping [6].

**Table 3A.** Estimated carrier frequency comparison

| Gene | Disease Name | Carrier frequency |  |  |
| --- | --- | --- | --- | --- |
|  |  | Our cohort | Westemeyer et al.[31] | C.-Y.Wei et al.[6] |
| *GJB2* | Hearing Loss(non-syndromic) | 16.68% | 6.25% | 1.59% |
| *VPS13B* | Cohen Syndrome | 2.78% | 0.33% |  |
| *CFTR* | Cystic Fibrosis* | 2.73% | 3.85% |  |
| *GALT* | Galactosemia | 2.64% | 0.67% |  |
| *SLC25A13* | Citrin Deficiency | 2.57% | 0.40% | 1.94% |
| *DUOX2* | congenital hypothyroidism | 2.34% |  |  |
| *GALC* | Krabbe Disease | 2.18% | 1.27% | 1.67% |
| *SLC26A4* | Pendred Syndrome | 2.04% | 1.43% | 1.70% |
| *ATP7B* | Wilson Disease | 1.98% | 1.52% | 1.77% |
| *G6PD* | G6PD deficiency | 1.98% |  | 2.49% |
| *PAH* | Phenylketonuria | 1.32% | 2.50% | 0.48% |
| *HBB* | Beta-Hemoglobinopathies | 1.32% | 3.23% | 0.59% |
| *PTS* | PTPS Deficiency(PKU) | 0.66% | 0.12% |  |

\*rs551227135(carrier frequency 1.9%) is so called *CFTR* IVS8-5T variant in previous literature. It is considered a “mild” *CFTR* mutation. When in trans with a known pathogenic *CFTR* mutation (e.g. $\Delta F508$ ), the IVS8-5T allele is responsible for non-classic CF phenotype, such as bilateral absence of vas deferens, recurrent pancreatitis, and late onset cystic fibrosis.[44] Individuals homozygous for the IVS8-5T allele as the sole variation of the whole *CFTR* coding sequence may present as non-classic CF with sinopulmonary disease and male infertility in Taiwan.[45]

Another example is the *SLC25A13* gene, related to citrullinemia. Pathogenic/likely pathogenic carrier frequency is 2.57% in our series, whereas it is only 0.40% in the U.S. cohort [31], and 1.9% in the TWBv2 series [6]. Yet another one is the *PTS* gene, defective *PTS* variants would lead to phenylketonuria. The pathogenic/likely pathogenic carrier frequency of *PTS* is 0.66% in our cohort, which is only 0.12% in the U.S. counterpart [31]. The frequency is compatible with clinical observation of PKU patients in Taiwan, where BH4-deficiency(defective *PTS*) PKU patients account for up to 1/4 of total PKU patients. In contrast, defective *PTS* PKU patients only account for 1-2% Caucasian cohorts. *PTS* variant is not shown in TWBv2 array data.

Based on TWB 1496 NGS cohort data, we applied the same method used in Westemeyer et al.[31] to calculate the combined at-risk couple rate in Taiwan. The 270 gene panel (274 genes in Westemeyer et al.[31]), omit 4 genes *HBA1/2, DMD, SMN1, FMR1*) would identify the risk for a genetic disorder in the offspring of 1 in 28 couples (3.55%). If *DUOX2*, *G6PD* were added to the local carrier screening list, the risk identification would be 1 in 25 couples (3.94%). (Table 3B)

**Table 3B.** Reproductive risk detection rate with different panels

| Expanded carrier gene panel | accumulated risk |  |
| --- | --- | --- |
| 270 gene panel | 3.55% | 1 in 28 couples at risk |
| 270 gene panel+*DUOX2*+*G6PD* | 3.94% | 1 in 25 couples at risk |
| 272 gene panel-adjustment(*CFTR*/*GALT*)* | 3.84% | 1 in 26 couples at risk |
| 272 gene panel-adjustment(*CFTR*/*GALT*/*GJB2*)* | 1.17% | 1 in 84 couples at risk |

\*after elimination of *CFTR*(rs551227135) / *GALT*(rs2070074) / *GJB2*(rs72474224) homozygous weak pathogenic variant condition

For the highest SV related hereditary disease in Taiwan: thalassemia, we used Manta [23] and AnnotSV [24] to identify *HBA1/2* pathogenic variants and in a subset of the TWB cohort

(N=494). The alpha thalassemia carrier frequency was 6.88%(5.06% α^0^, 1.81% α^+^)(Table 4A). For the other high prevalence hereditary disease spinal muscular atrophy (SMA), the *SMN1/SMN2* copy number was analyzed by SMNCopyNumberCaller[10]. 10 carriers with only one copy of *SMN1* were identified out of 494 members, which meant 2.02% carrier frequency of SMA(Table 4B). Validation of NGS based SMA carrier screening methods in China [32] [33] convinced us that these are as accurate as the traditional MLPA method. The comparison of alpha thalassemia carrier and SMA carrier rate in neighboring Asian regions are listed in Table S4 and S5.

**Table 4A.** Analysis of 494-people subset of HBA1/2 carrier status(using Manta and AnnotSV)

| Thalassemia carrier | $\alpha^+$ | | | | $\alpha^0$ | | |
| --- | --- | --- | --- | --- | --- | --- | --- |
| Genotype | $-\alpha^{3.7}$ | $\alpha^{CS}$<br>(rs41464951) | $\alpha^{QS}$<br>(rs41397847) | $\alpha^{WS}$<br>(rs41479347) | --SEA | --THAI | --FIL |
| N= 494 | 2 | 1 | 1 | 5 | 22 | 2 | 1 |

**Table 4B.** The *SMN1/SMN2* copy number (CN) distribution from 494 Taiwanese.

|  | *SMN1* CN |  |  |  |  |  |
| --- | --- | --- | --- | --- | --- | --- |
| *SMN2* CN |  | **0**<br>(SMA patient) | **1<sup>∞</sup>**<br>(SMA carrier) | **2** | **3** | **4** |
|  | **0** | 0 | 0 | 21 | 5 | 0 |
|  | **1** | 0 | 0 | 145 | 18 | 1 |
|  | **2** | 0 | 4 | 278 | 4 | 0 |
|  | **3** | 0 | 6 | 9 | 2 | 0 |
|  | **4** | 0 | 0 | 0 | 0 | 0 |

¶ SMNCopyNumberCaller fails to identify *SMN1/SMN2* status in 1 of 494 individuals
∞There are 10 carriers out of 494 cases. 4 of them are *SMN1*:1/*SMN2*:2; 6 of them are *SMN1*:1/*SMN2*:3.

### Variants with clinical significance and high allele frequency in Taiwan

Initial data filtering with allele frequencies singled out 54 variants with MAF greater than

0.01 and 24 with MAF between 0.005 and 0.01 among the carrier call-set from 1,496 TWB participants. To address the characteristic of Taiwanese population, we further filtered the 78 variants by comparing their MAF in the gnomAD genome database (version 2.1.1) and the ExAC database. The results single out 15 variants with clinical significance and relatively high MAF in Taiwan (Table S3).

Variants with MAF ≧ 0.01 among Taiwan Biobank participants

### Splice-donor variant in CACNA1B

A splice-donor variant in CACNA1B c.390+1_390+2insACGACACGGAGCCCTATTTCATCGGGATCTTTTGCTTCGAGGCAG GGATCAAAATCA occurs with a MAF of 0.019 among 1,496 TWB participants, while this variant is not reported in ExAc and the East Asian population of gnomAD.

### Stop-gained variant in DUOX2

Stop-gained variant c.1588A>T in DUOX2 occurs with a MAF of 0.013 among TWB participants, while its MAF in the East Asian population in the gnomAD database is 0.0071. The protein product of DUOX2 is an oxidase and part of the peroxide-generating system located at the apical membrane of thyroid follicular cells [34, 35]. Prematurely terminated protein products of DUOX2 with the pathogenic variant we herein mentioned lead to a lower level of hydrogen peroxide and consequently insufficient thyroid hormone needed for normal human development [36]. This variant has been identified in patients with transient or permanent hypothyroidism as well as iodide organification [36, 37].

### Frameshift variant in VPS33A

A frameshift variant in VPS33A c.13dup occurs with a MAF of 0.012 among 1,496 TWB participants, while its MAF in the East Asian population of gnomAD is 0.0096.

Variants with 0.01≧ MAF > 0.005 among Taiwan Biobank participants

### Missense variant in OPN1MW

Missense variant c.989G>A in OPN1MW is reported to be causative of deuteranopia. OPN1MW, the medium-wave-sensitive opsin-1 gene, is mapped to chromosome Xq28 and encodes the green cone pigment, which is key to color vision. The absorbance of the Arg330Gln mutant opsin decreased dramatically compared to normal green opsin. The presence of c.989G>A variant in OPN1MW has been reported to be causative of deutan color blindness [38]. A precise study on the prevalence of deuteranopia in Taiwan may help explain the relatively high allele frequency of c.989G>A variant among Taiwan Biobank participants (MAF = 0.008) in OPN1MW compared to that in gnomAD East Asian population.

### Intronic variants in SLC25A13

A variant, c.615+5G>A in SLC25A13 occurs with a MAF of 0.006 in TWB participants. SLC25A13, localized to chromosome 7q21.3, encodes citrin, which serves as a mitochondrial solute transporter in the urea cycle. c.615+5G>A are linked to neonatal intrahepatic cholestasis caused by citrin deficiency (NICCD) and adult-onset type II citrullinemia (CTLN2) [39–41]

### Frameshift variant in SERPINB7

Splice-acceptor variant c.522dup in SERPINB7 occurs with a MAF of 0.005 among TWB participants, while its allele frequency in the East Asian population in the gnomAD2 database is 0.0032. This variant is causative of Nagashima-type palmoplantar keratoderma, a skin disorder characterized by hyperkeratosis of the palm and feet of affected individuals [42, 43]

### Frameshift variant in TTLL5

A frameshift variant in TTL5 c.3177_3180del occurs with a MAF of 0.005 among 1,496 TWB participants, while its MAF in ExAc and in the East Asian population of gnomAD is 0.0021 and 0.0045, respectively.

### TaiwanGenomes (genomes.tw)

TaiwanGenomes (https://genomes.tw) provides a user-friendly interface to users to access all the variants reported in this study. The ’MAIN’ tab links to the main table that contains 59,433,212 variants outputted by joint calling of the 1,496 WGS, including 51,135,411 variants passing the VQSR analysis (setting ‘VQSR = PASS’). On the other hand, the ’SUPPLEMENT’ tab links to the supplementary table that provides the information of read depth for 2,792,591,408 positions in the human genome. These positions are categorized into four classes: A: Ref (MAF=0), B: Missing (MAF=n.a), and C: Uncertain quality (MAF=n.a/0), and the variants called by GATK (59,433,212 variants) have links to the MAIN table. TaiwanGenomes provides users the flexibility of selecting columns of interest to examine. Among the PASS variants, 439,192 variants are falling in the coding regions (setting ‘FILTER=PASS’ and ‘fun.refGene=exonic’), and 55,949 of them have a minor allele frequency ≥0.01 (setting ‘FILTER=PASS’ and ‘fun.refGene=exonic’ and ‘AF≥0.01’).

Another example of setting condition combinations of multiple selected columns is: a user can examine all the non-synonymous variants in BRCA1 by setting ‘Gene.refGene=BRCA1’ and ‘ExonicFunc.refGene=nonsynonymous_SNV’, resulting in 40 variants.

## Discussion

Unlike the genotyping array that focuses on detecting the associations between genomic haplotype block and phenotypic traits, personal genome sequences directly uncover all the hereditary risks. In this study, we reanalyzed 1,496 whole-genome sequence data from the Taiwan Biobank, which is one of a few valuable datasets for the east Asian population. Our reanalyzed results disclosed the comprehensive medical genomic profile which is complement to previous Taiwan Biobank studies and demonstrated the potential of what WGS data can reveal. We utilized the benchmarked data from GIAB and jointly-called 1,496 samples to determine the cut-off VQSR tranche at 99.7, and provided VQSR quality flags for each locus on the website (**TaiwanGenomes** https://genomes.tw) . We further selected hundreds of variants without considering the VQSR tranche and validated them by the Sanger sequencing. Firstly, for the variants that were removed in the joint calling step, the results of sanger sequencing are all negative, revealing the effectiveness of joint calling to eliminate the false positives in individual variant calling. On the other hand, of all the variants that were rescued by joint calling, only some of them have positive results in Sanger sequencing, suggesting further VQSR filtering criteria is necessary. For the reference alleles across the genome, we defined all non-alternative allele regions into three categories by the overall call rate and the depth of coverage. Consequently, it is clear to distinguish the reference homozygous or the difficult-to-map regions in the genome reference for TWB 1496 cohort.

We found a taiwanese has on average 6871 globally novel variants compared to other existing databases including gnomAD.

In this study, we utilized WGS data to estimate the carrier risk in Taiwan. For the secondary finding, we analyzed the variant on ACMG SF v3.0 genes. This is the first time ACMG SF v3.0 has applied to the East Asian population. We found that 5.54% of the 1,496 cohort are carriers of ACMG SF v3.0 genes, 1.67% are autosomal dominant and X-linked diseases, 3.87% are recessive diseases. Notably, one pathogenic variant NM_000314.6:c.802-2A>T on the *PTEN* gene was indeed a technical error in TWB 1,496 cohort. The variant located at the end of a polyT region with strand bias evidence in our analysis, highlighting the necessity of applying VQSR and quality check in WGS data reanalysis. We also analyzed a 270 gene panel to estimate the accumulated monogenic disease risk and found 3.55% of offspring would be the carriers, implying approximately 1 of 28 couples might benefit from the carrier screening. Two main pathogenic variant contributors are *GJB2:*NP_003995.2:p.Val37Ile (rs72474224, MAF:8.6%) and *CFTR:*NM_000492.4:c.1210-11_1210-10insG (rs551227135, MAF:0.9%). Both are considered “mild” pathogenic variants. Phenotype penetration of *GJB2*(rs72474224) is highly variable in Taiwan clinical experience. *CFTR*(rs551227135) is one of the classical *CFTR* IVS8-5T variants, which are responsible for non-classical cystic fibrosis presentation(CABVD, recurrent pancreatitis, late-onset CF). The regional normalization of variants is complicated and annotations are still controversial. Besides, another caveat for carrier screening is the inability to detect long fragment variants from short-reads data, e.g. CNV, large deletion, trinucleotide repeats, and gene conversion. With the short-reads data, we often underestimate the carrier frequency of 4 important genes: *HBA1/2*, *DMD*, *FMR1*, *SMN1*, of which many known pathogenic variants are relatively large.

With regard to long-fragment genetic variants, we randomly selected 494 samples for deciphering the population structure variation profile on thalassemia genes *HBA1/2* and the copy number on *SMN1/2* genes. The overall population profile was similar to previous studies by target panel approach, suggesting that WGS can potentially replace target panel once the analysis pipeline has been built (supplementary table) Furthermore, our results showed 39.8% (439/1103) of the cohort was vulnerable to severe ADRs since they carry at least one risk *HLA* allele. Even though several studies implied the HLA types could be inferred by SNP genotyping results, full HLA haplotypes from sequence data was unbiased for population specific rare haplotypes. Similarly, SNVs on CYP genes could be detected by SNP genotyping, but WGS can reveal completed haplotype information which is more accurate providing susceptibility. Our data revealed Taiwanese carry high frequency (MAF > 10%) abnormal alleles in more than 60% of the pharmacogenetic variant-drug groups. It is also noteworthy that TWB subjects were 20 years-old adults with no distinct developmental defects or malignant tumor at the times when they joined the study.

## Conclusions

The WGS has become affordable for the research and clinic. As a person only needs to test once for a lifetime, more comprehensive reanalysis of genomic profile will have great clinical impact. Overall, our study highlights the potential of a complete genomic profile with medical information for a population and individuals.

## Data Availability

The WGS data from 1,496 TWB participants are available through the TWB (https://www.twbiobank.org.tw/new_web_en/about-export.php).
The allele frequency data are available online at (https://genomes.tw/#/).

---

**Figure S1.**
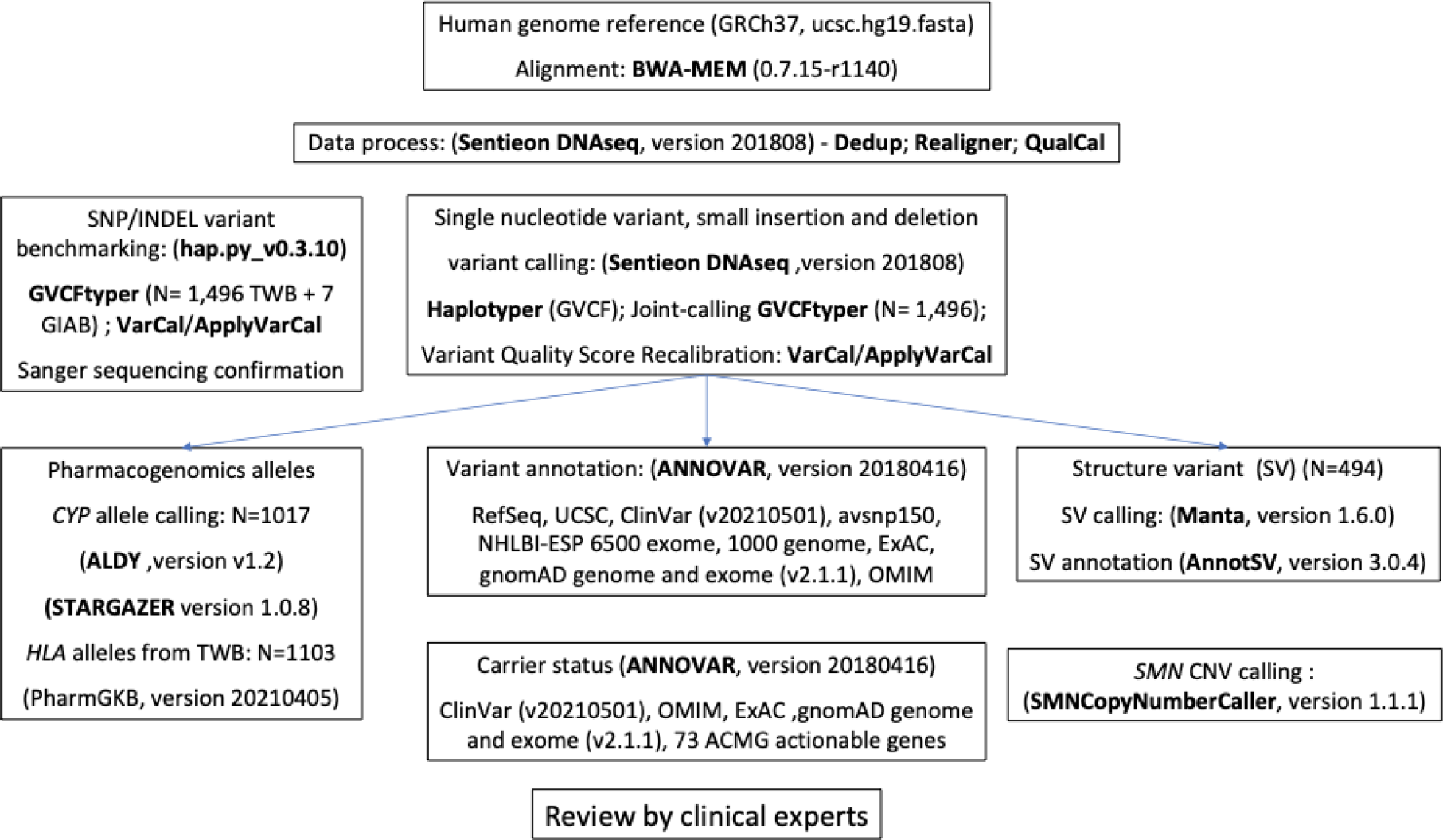
The overall study design and analysis pipelines. Software names and functions are shown in bold text.

**Figure S2.**
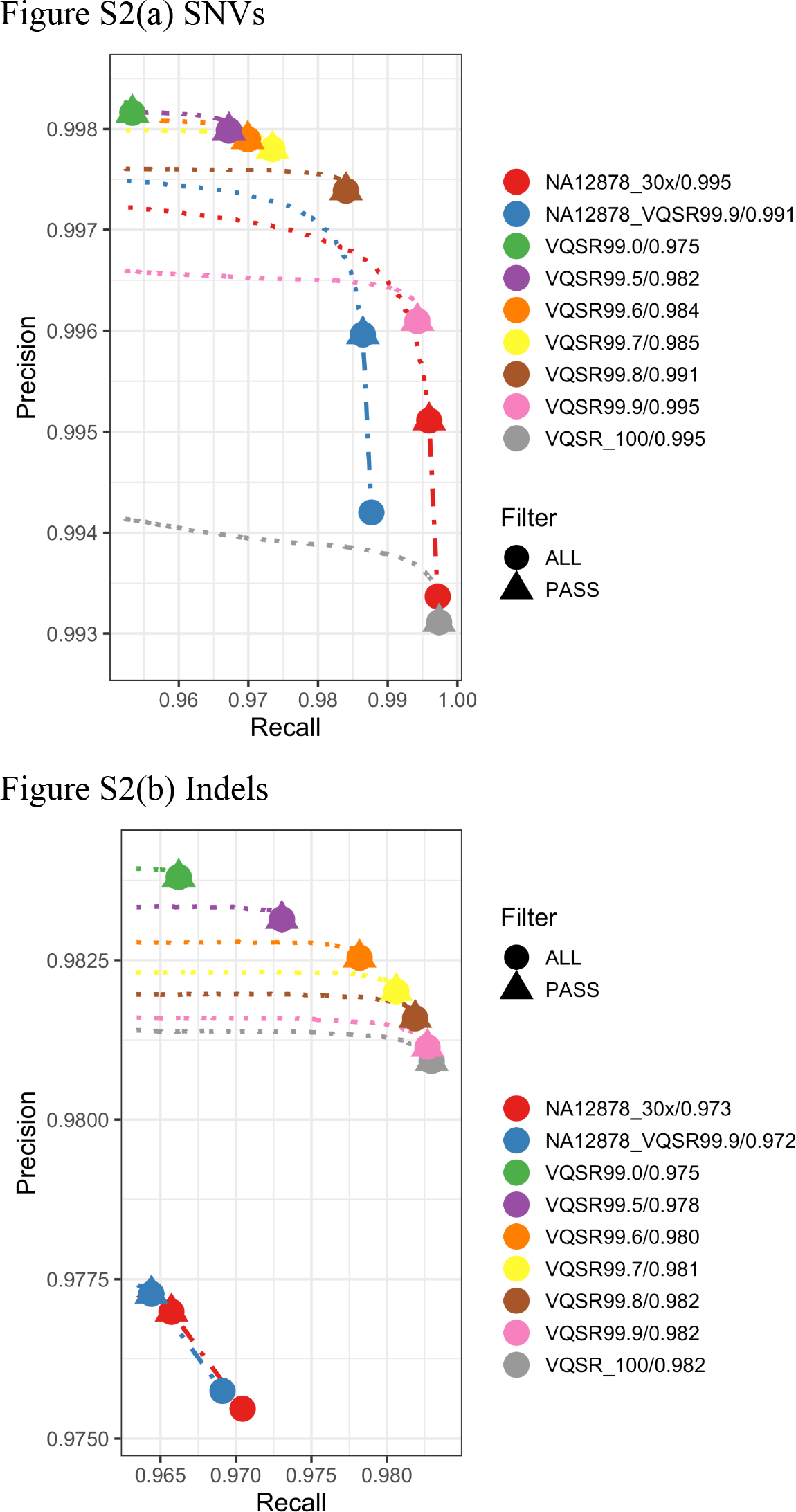
Precision-Recall (PR) curves given by seven variant filtering cut-offs based on variant quality score recalibration algorithm for SNVs and INDELs. Comparison of overall accuracy and F1 scores are included. The circles depict the accuracy given by all variants under VQSR filtering cut-offs, and the triangles represent the accuracy given the variants pass Haplotyper default quality criteria. The red and blue indicate the performance given by single NA12878 data with the same pipeline.

**Figure S3A.**
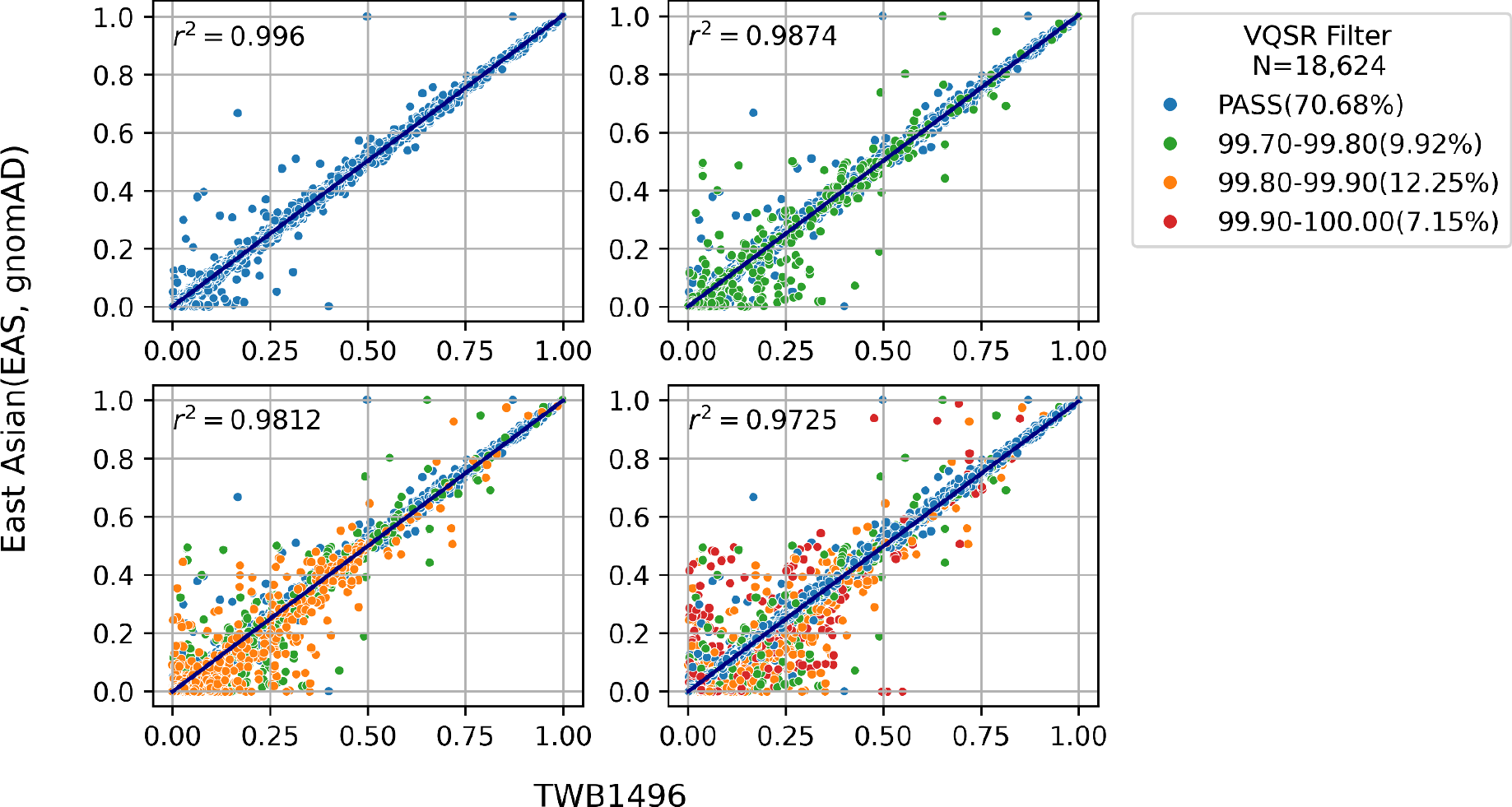
(Top-left), S3B(Top-right), S3C(Bottom-left), S3D(Bottom-right) Scatter plots to compare the minor allele frequency (MAF) of TWB1496 against gnomAD East Asian (EAS) v2.1 release. Four figures represent the correlation of MAF among different variant quality scores. Regression lines and r-squared are shown. X-axis: The MAF of 1496 Taiwan Biobank whole-genome sequencing samples. Y-axis: The MAF of East Asian data in gnomAD v2.1. Blue dots represent nonsynonymous variants whose VQSR score is PASS.

**Table S1.** Results of Sanger sequencing

|  | NGS2015051<br>0B |  | NGS2015051<br>0C |  | NGS2015051<br>0D |  | NGS2015051<br>0F |  | All sample |  |
| --- | --- | --- | --- | --- | --- | --- | --- | --- | --- | --- |
|  | ALL | PASS | ALL | PASS | ALL | PASS | ALL | PASS | ALL | PASS |
| True positive | 2 | 2 | 2 | 2 | 7 | 6 | 6 | 6 | 17 | 16 |
| False positive | 13 | 4 | 13 | 4 | 11 | 4 | 14 | 4 | 51 | 16 |
| True negative | 5 | 0 | 5 | 0 | 15 | 4 | 16 | 4 | 41 | 8 |
| FDR |  |  |  |  |  |  |  |  | 0.75 | 0.5 |
| TNR |  |  |  |  |  |  |  |  | 0.45 | 0.33 |

**Table S2A.** HLA variant-drug pairs and allele frequencies in the sample of 1,103 TWB volunteers.

| Haplotype | Gene | Allele frequency in TWB (N≤1103) | Level of evidence | Chemicals | Phenotypes |
| --- | --- | --- | --- | --- | --- |
| *HLA-A\*31:01:02* | *HLA-A* | 2.68% (59/2204)<sup>+</sup> | 1A | carbamazepine | Drug Hypersensitivity, drug reaction with eosinophilia and systemic symptoms, Epidermal Necrolysis, Toxic, Maculopapular Exanthema, Stevens-Johnson Syndrome |
| *HLA-B\*58:01* | *HLA-B* | 12.1% (267/2206)<sup>+</sup> | 1A | allopurinol | Drug Hypersensitivity, drug reaction with eosinophilia and systemic symptoms, Epidermal Necrolysis, Toxic, severe cutaneous adverse reactions, Stevens-Johnson Syndrome |
| *HLA-B\*15:02:01* | *HLA-B* | 3.72% (82/2206) | 1A | oxcarbazepine | Stevens-Johnson Syndrome |
| *HLA-B\*15:02:01* | *HLA-B* | 3.72% (82/2206) | 1A | phenytoin | drug reaction with eosinophilia and systemic symptoms, Epidermal Necrolysis, Toxic, severe cutaneous adverse reactions, Stevens-Johnson Syndrome |
| *HLA-B\*15:02:01* | *HLA-B* | 3.72% (82/2206) | 1A | carbamazepine | Epidermal Necrolysis, Toxic, severe cutaneous adverse reactions, Stevens-Johnson Syndrome |
| *HLA-B\*57:01:01* | *HLA-B* | 0.59% (13/2206) | 1A | flucloxacillin | drug-induced liver injury |
| *HLA-B\*57:01:01* | *HLA-B* | 0.59% (13/2206) | 1A | abacavir | Drug Hypersensitivity,<br>HIV Infections |
| *HLA-B\*40:01:01* | *HLA-B* | 18.81% (415/2206)<sup>s</sup> | 2A | carbamazepine | Epidermal Necrolysis, Toxic,<br>Stevens-Johnson Syndrome |
| *HLA-B\*13:01:01* | *HLA-B* | 5.58%(123/2206) | 2A | dapsone | Drug Hypersensitivity, drug reaction with<br>eosinophilia and systemic symptoms,<br>Leprosy, Maculopapular Exanthema,<br>severe cutaneous adverse reactions,<br>Stevens-Johnson Syndrome |
| *HLA-B\*15:11:01* | *HLA-B* | 1.04% (23/2206) | 2A | carbamazepine | severe cutaneous adverse reactions |
| *HLA-B\*59:01:01:01* | *HLA-B* | NA | 2A | methazolamide | Epidermal Necrolysis, Toxic,<br>Stevens-Johnson Syndrome |
| *HLA-A\*33:03* | *HLA-A* | 12.93% (285/2204)<sup>+</sup> | 2B | allopurinol | Drug Hypersensitivity,<br>Stevens-Johnson Syndrome |
| *HLA-C\*01:02:01* | *HLA-C* | 16.05% (354/2206) | 2B | methazolamide | Epidermal Necrolysis, Toxic,<br>Stevens-Johnson Syndrome |
| *HLA-C\*03:02* | *HLA-C* | 12.15% (268/2206)<sup>+</sup> | 2B | allopurinol | Epidermal Necrolysis, Toxic, severe cutaneous adverse reactions, Stevens-Johnson Syndrome |
| *HLA-C\*04:01:01:01* | *HLA-C* | 3.99% (88/2206) | 2B | nevirapine | drug reaction with eosinophilia and systemic symptoms, Epidermal Necrolysis, Toxic, Exanthema, Hypersensitivity, Stevens-Johnson Syndrome |
| *HLA-DPB1\*03:01:01* | *HLA-DPB1* | 5.29% (109/2059) | 2B | aspirin | Asthma |
| *HLA-DRB1\*01:01:01* | *HLA-DRB1* | 0.77% (17/2206) | 2B | nevirapine |  |

HLA: human leukocyte antigen; TWB: Taiwan biobank; (\$) denoted HLA-B\*40:01:01 has zero AF, whereas HLA-B\*40:01:02 and HLA-B\*40:01:45 alleles with identical CDS sequences are common in Taiwanese population. (+) denoted TWB database released more nomenclature fields than PharmGKB listed fields.

**Table S2B.**
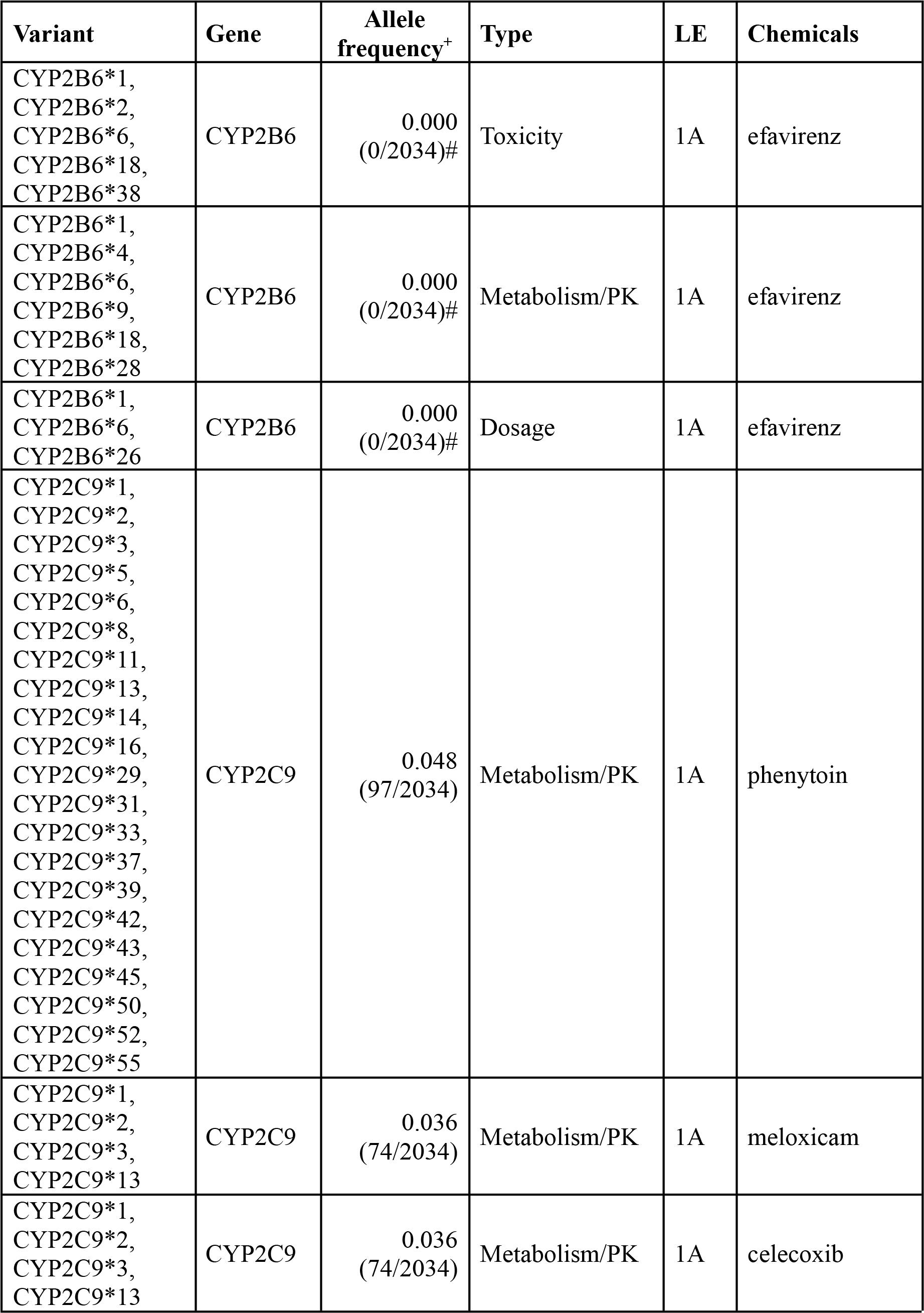
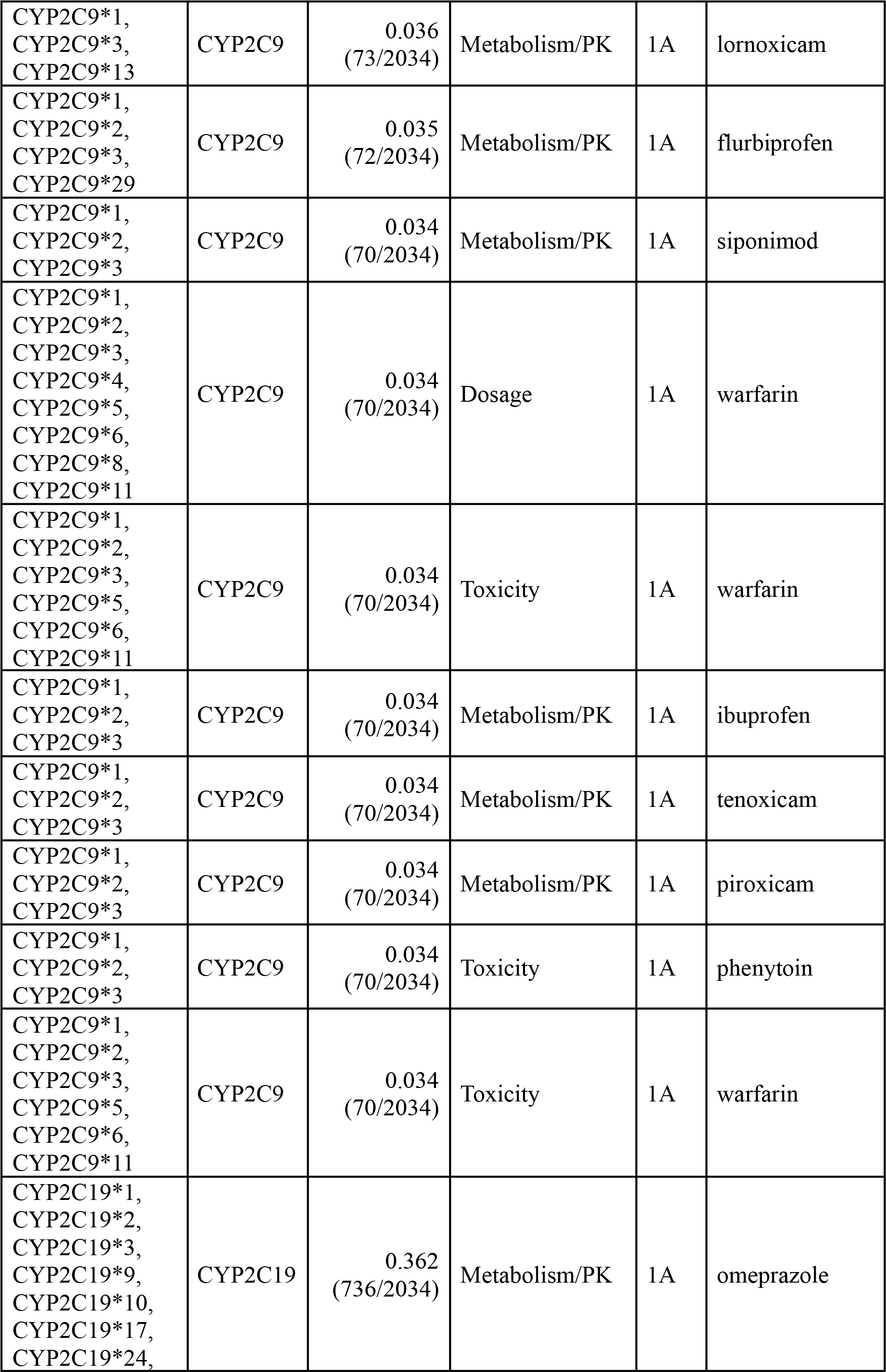
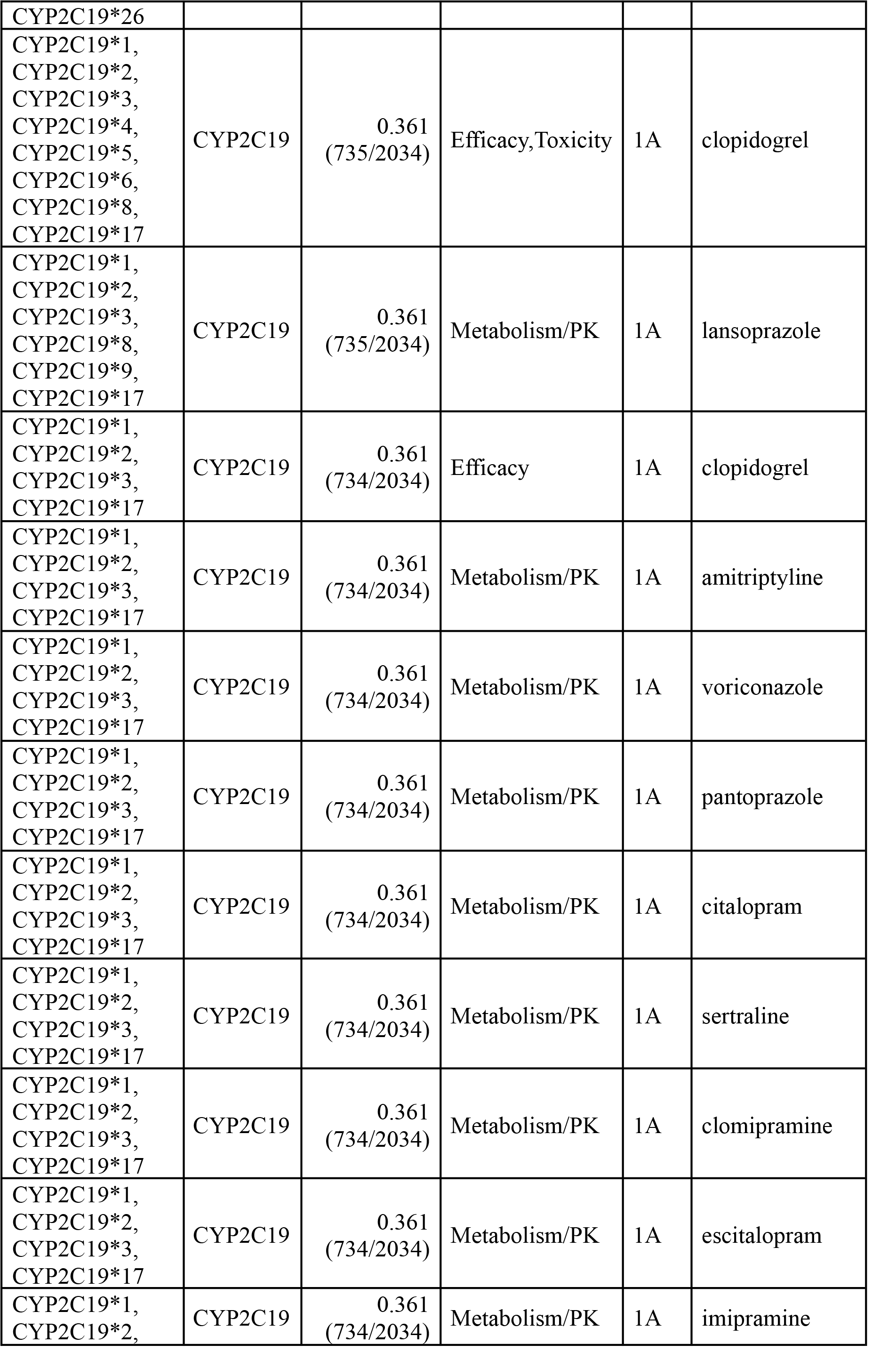
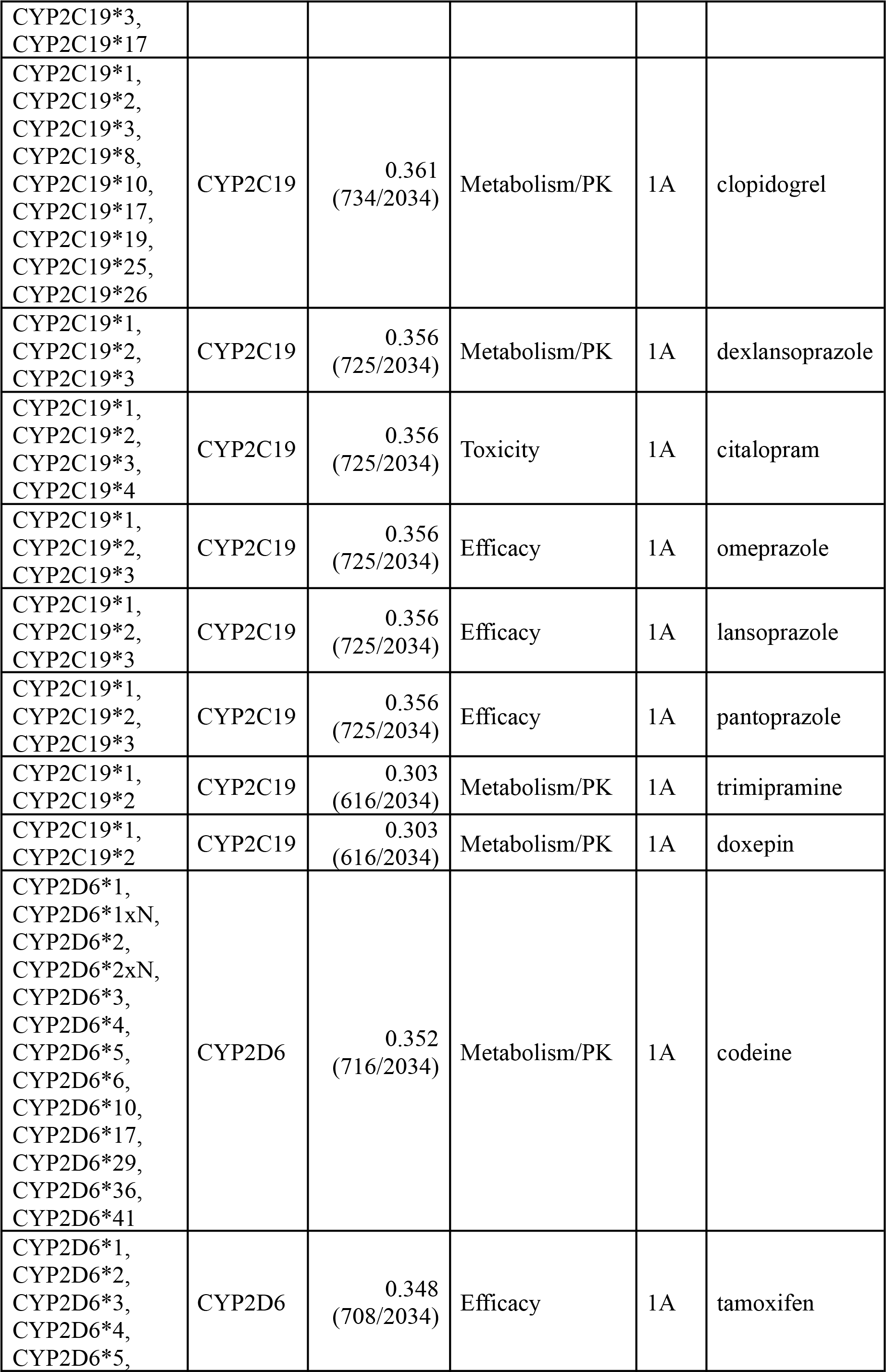
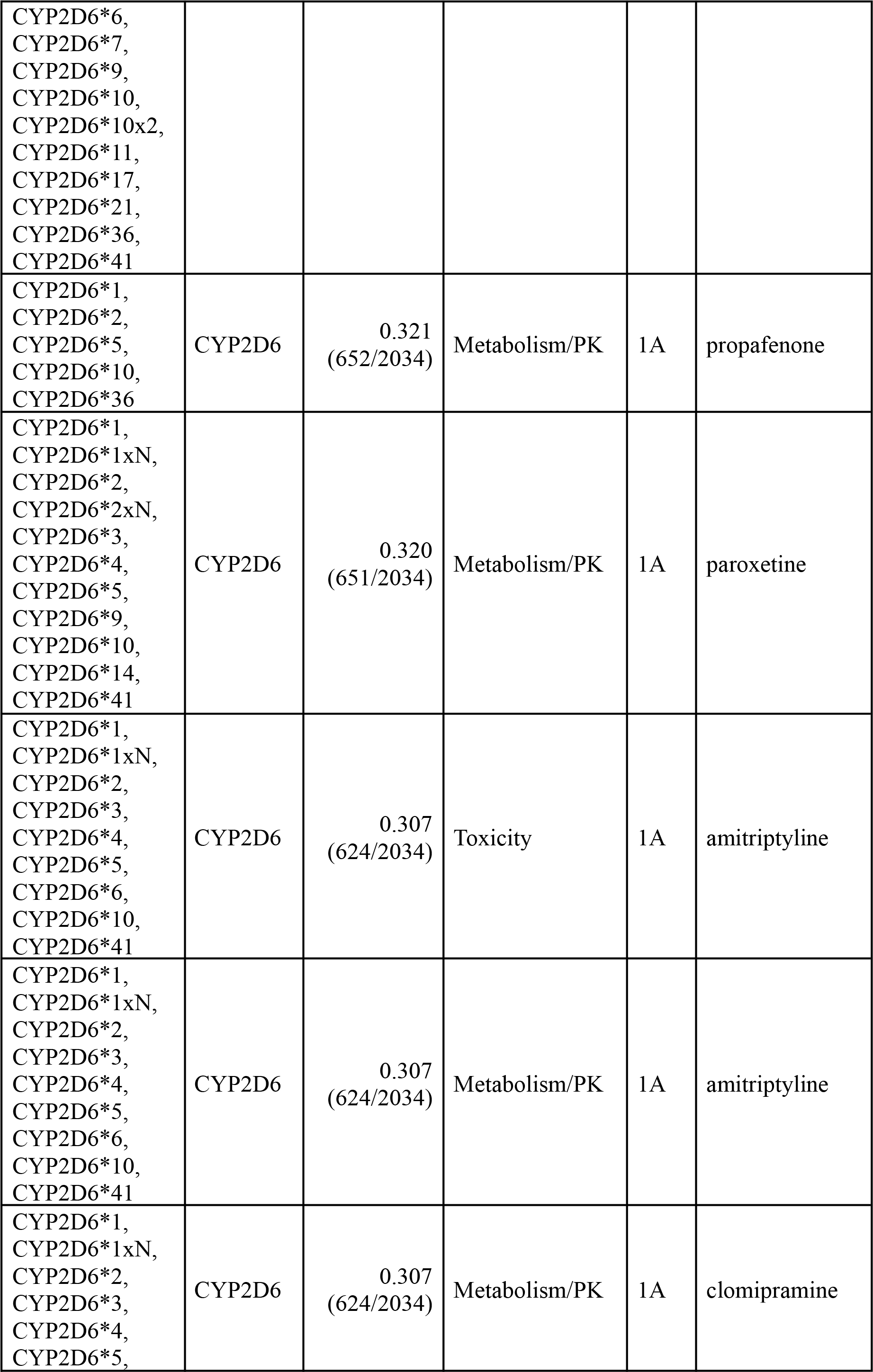
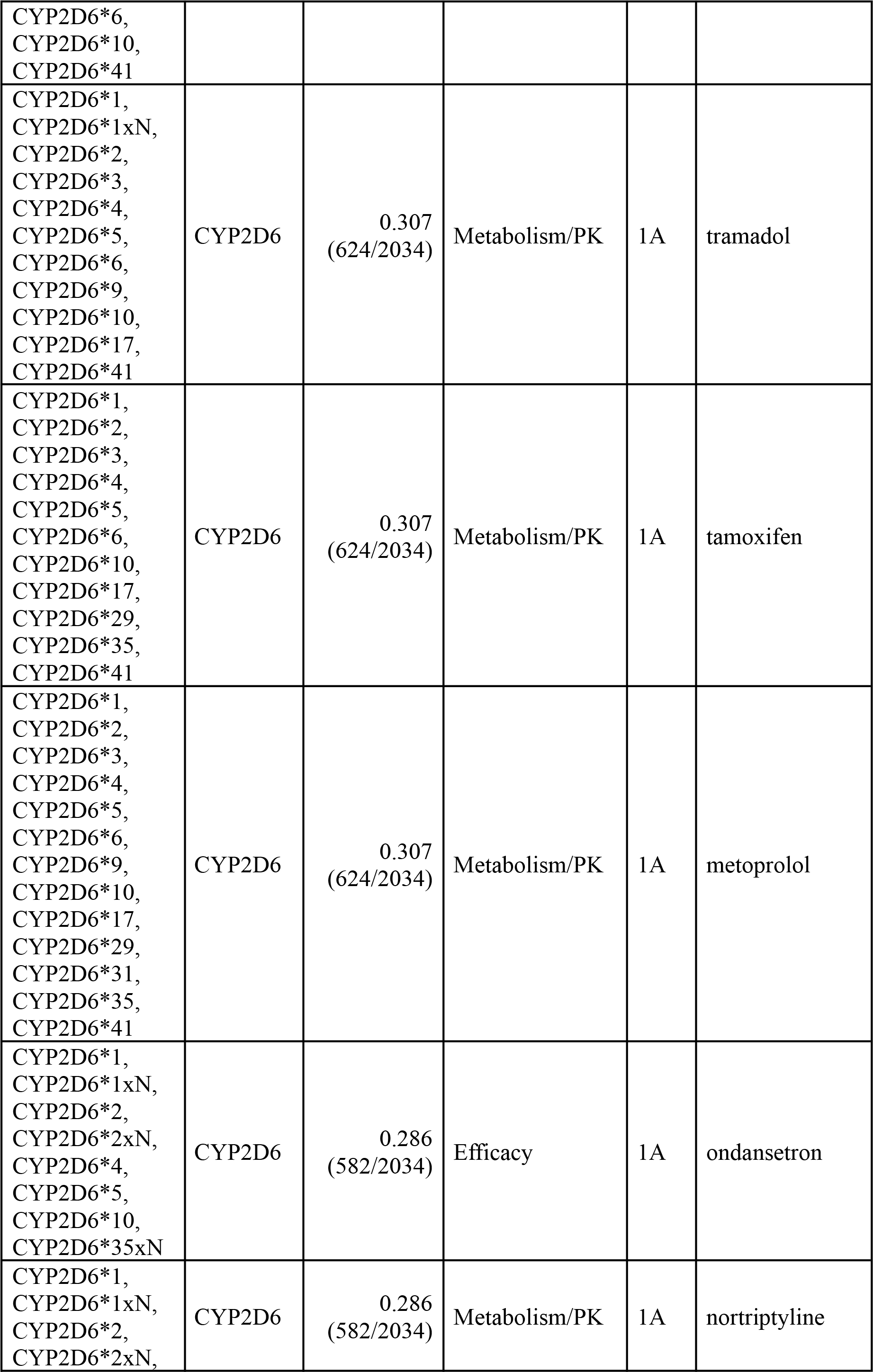
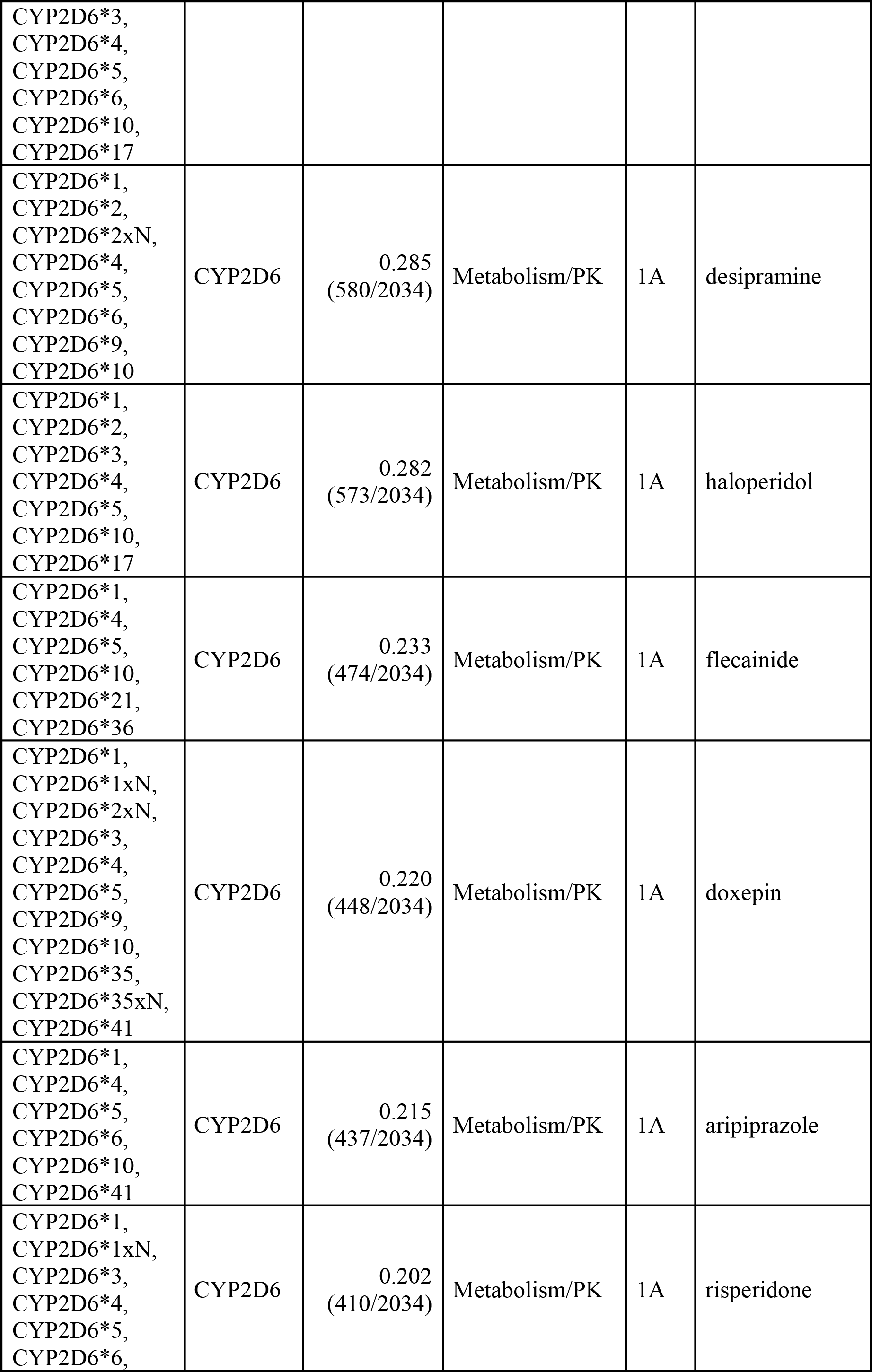
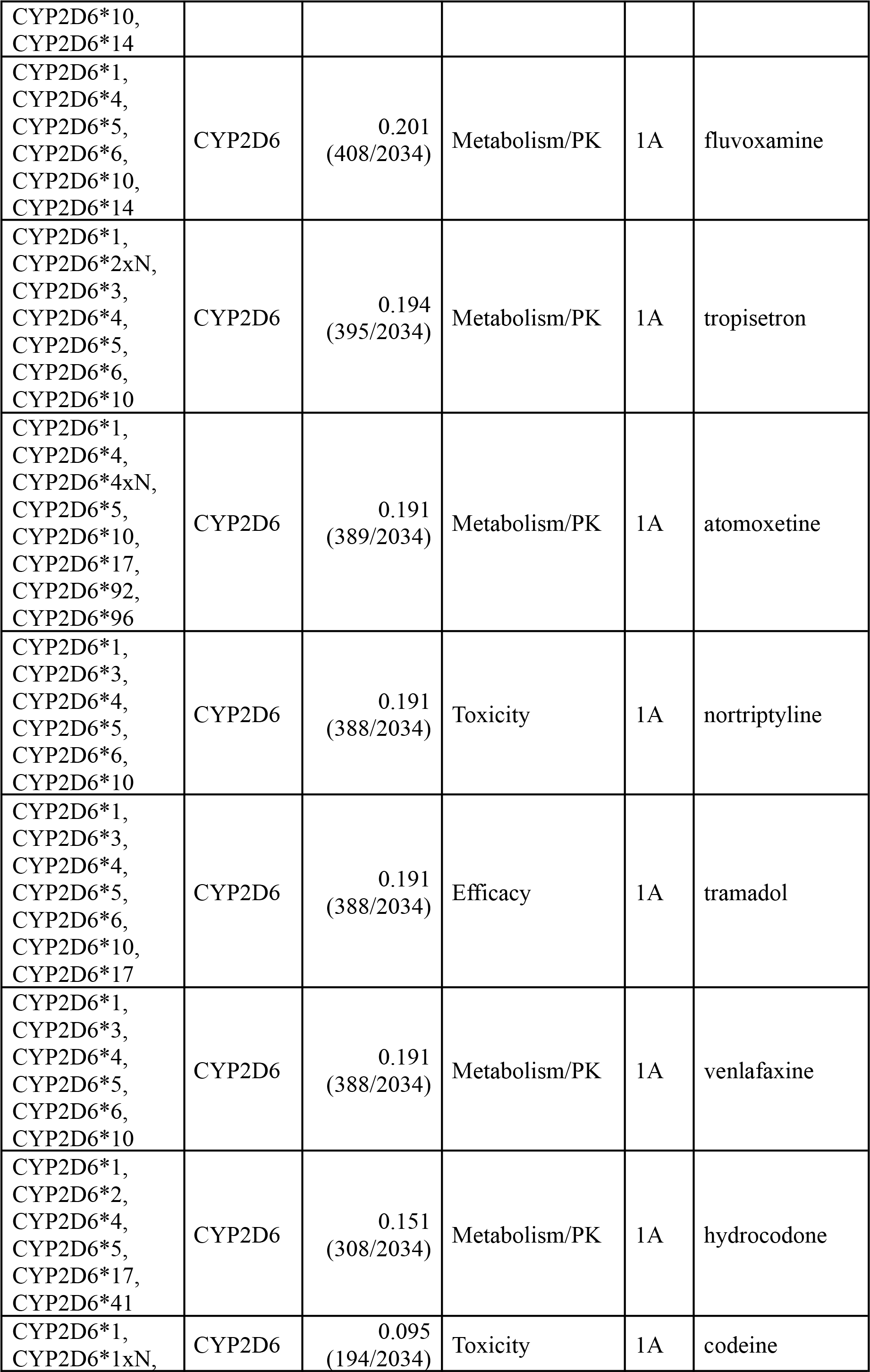
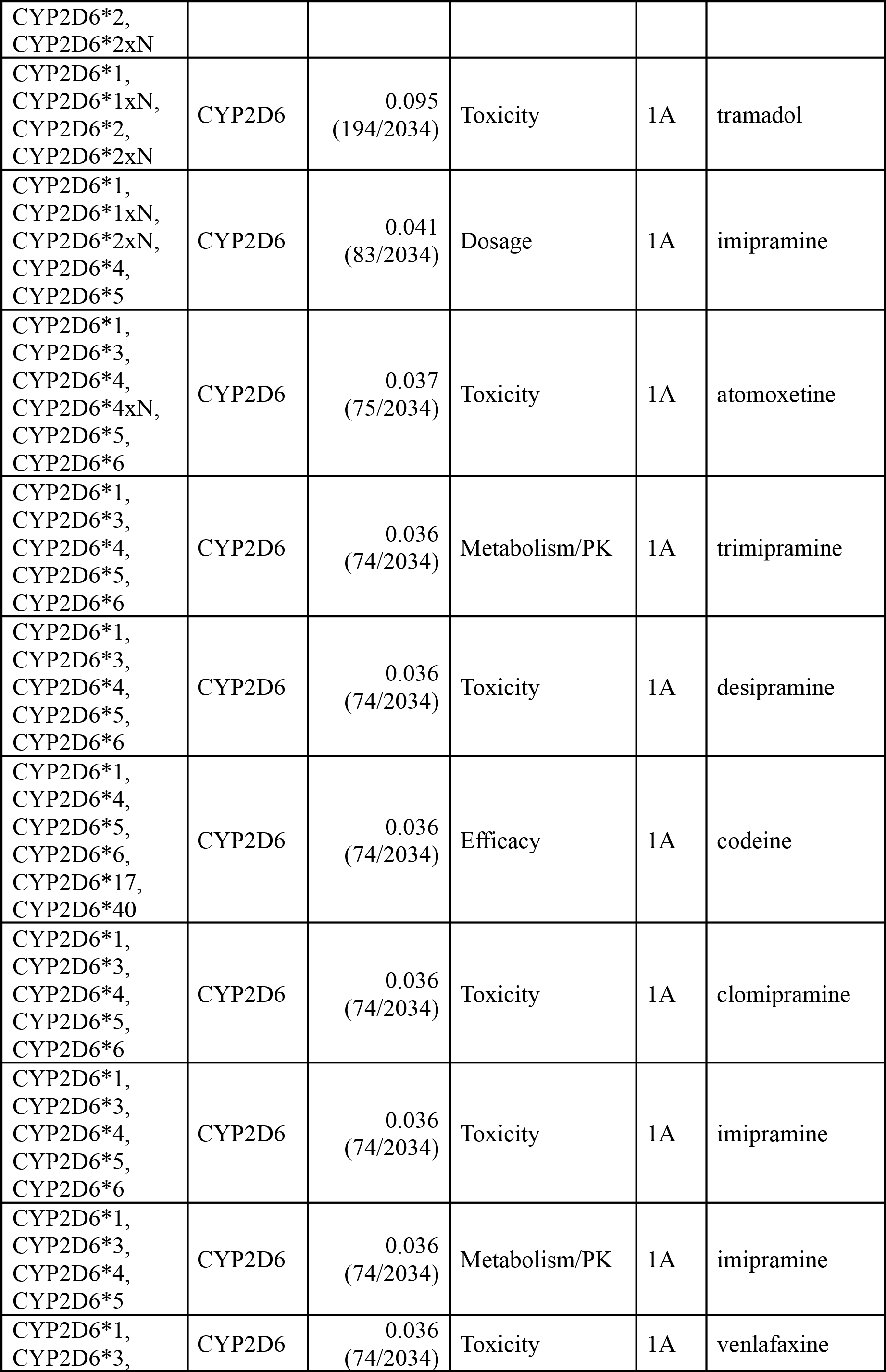
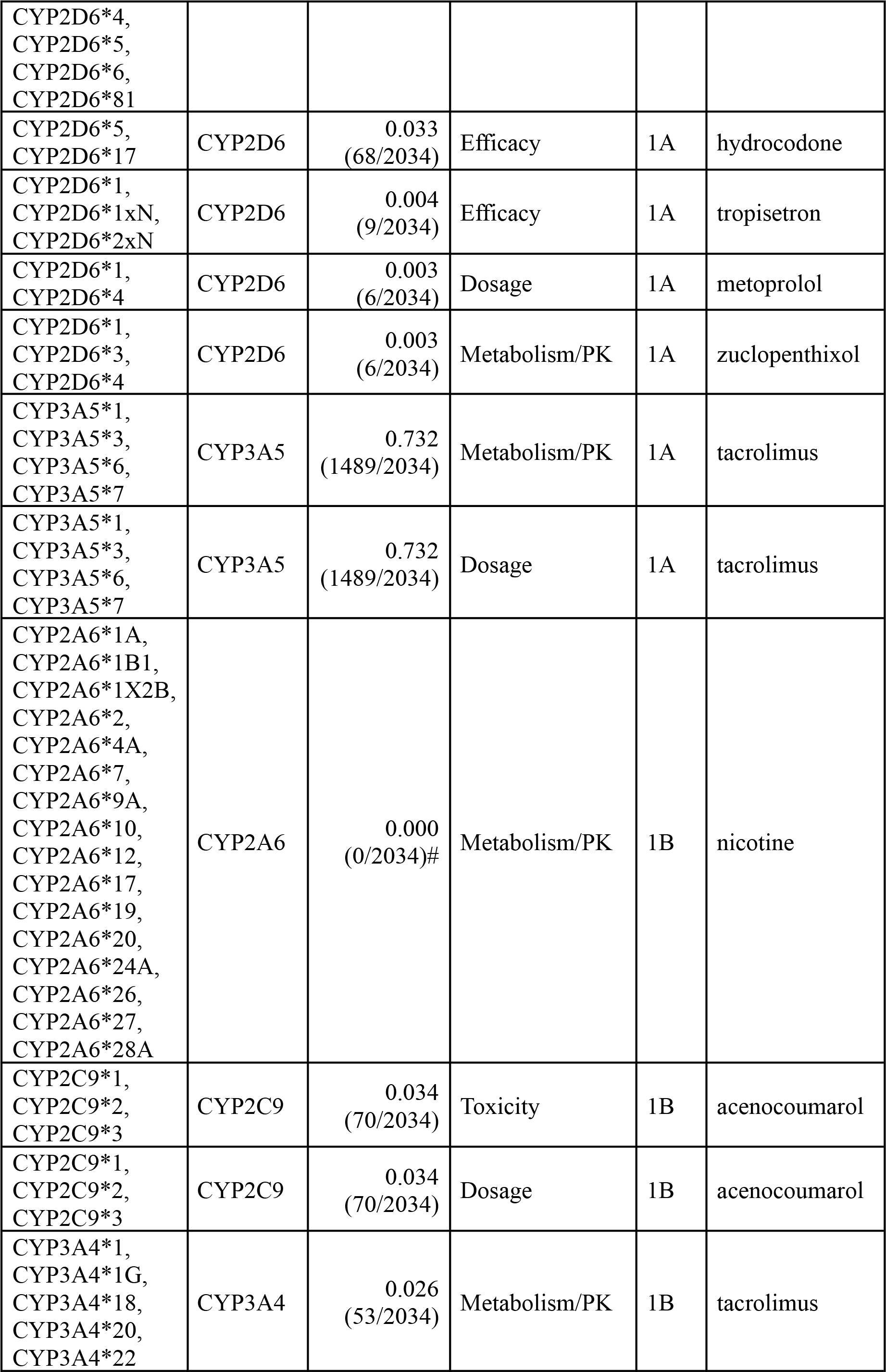
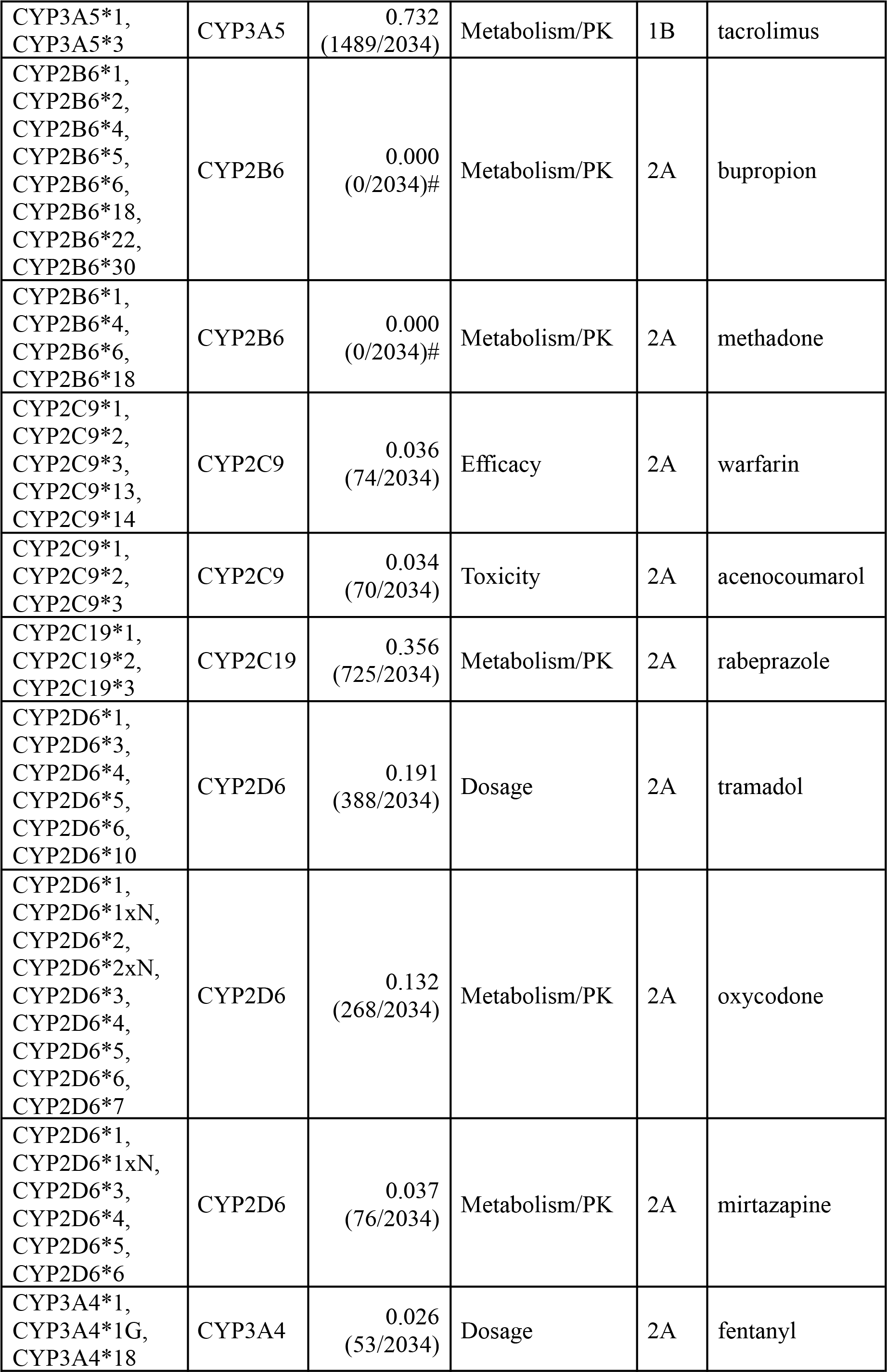
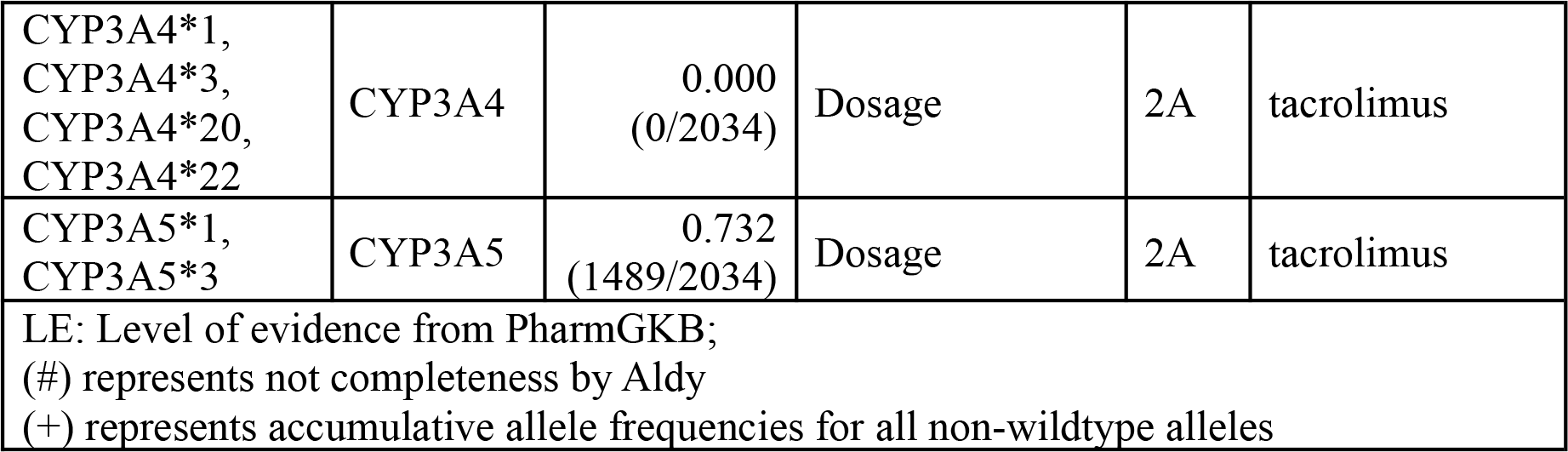
*CYP* variant-drug pairs and allele frequencies in the sample of 1,017 TWB volunteers.

**Table S3.**
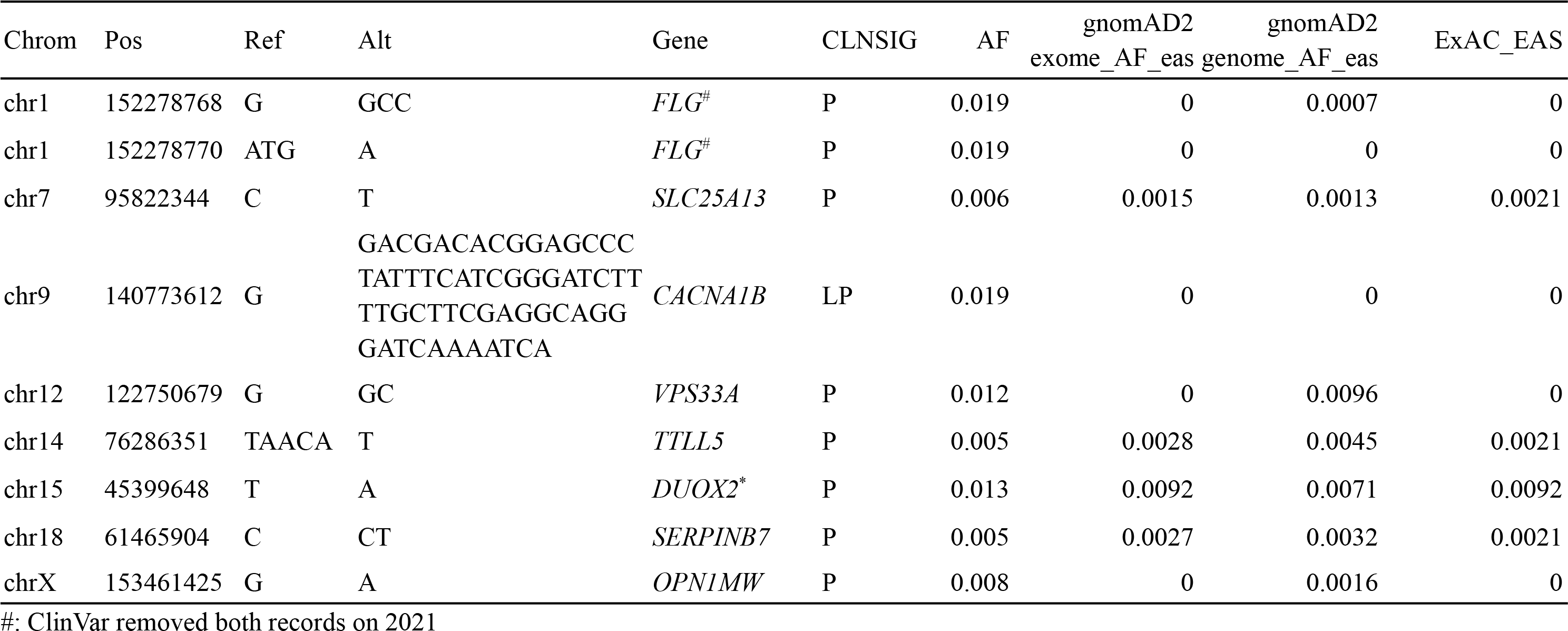
Variants with clinical significance and high allele frequency in Taiwan Biobank

**Table S4.** Comparison of alpha thalassemia carrier rate in Southeastern Asia

|  |  |  |  |  |
| --- | --- | --- | --- | --- |
|  | Taiwan biobank<br>(n=494) | China<br>(Fujian, n=189,414)[46] | China<br>(Guangdong, n=4,622)[9] | China<br>(Dai nationality in Yunnan, n=951)[47] |
| Detection method | NGS analysis | MCV<80 and HbA2<2.5% | NGS analysis | 1) MCV<80 and HbA2<2.5%<br>2) NGS analysis |
| Alpha thalassemia Carrier frequency | 6.88% | 4.84% | 12.7% | 29.9%(blood)<br>39.1%(NGS) |

**Table S5.** Comparison of SMA carrier frequency in Asia

|  | Taiwan biobank<br>(n=494) | China<br>(N=13069)[48] | Thailand<br>(N=505)[49] | Korea<br>(N=1581)[50] |
| --- | --- | --- | --- | --- |
| Detection method | NGS & SMNCopyNumber Caller | MLPA | MLPA | MLPA |
| SMA carrier frequency | 2.02% | 1.77% | 1.78% | 1.83% |

## Notes

### Competing Interest Statement

The authors have declared no competing interest.

### Funding Statement

This study was funded by the Ministry of Science and Technology, Taiwan (MOST 109-2622-B-002-004-CC2, MOST 109-2221-E-002-162-MY3 and MOST 110-2320-B-002-078-) and National Taiwan University Hospital (UN109-070).

### Author Declarations

The IRB on Biomedical Science Research of Academia Sinica, Taiwan (IRB-BM) and the Ethics and Governance Council (EGC) of Taiwan Biobank, Taiwan gave ethical approval for this work.

## References

1. Stark Z, Dolman L, Manolio TA, Ozenberger B, Hill SL, Caulfied MJ, et al. Integrating Genomics into Healthcare: A Global Responsibility. Am J Hum Genet. Elsevier; 2019;104:13–20.

2. Tadaka S, Katsuoka F, Ueki M, Kojima K, Makino S, Saito S, et al. 3.5KJPNv2: an allele frequency panel of 3552 Japanese individuals including the X chromosome. Hum Genome Var. 2019;6:28.

3. Okada Y, Momozawa Y, Sakaue S, Kanai M, Ishigaki K, Akiyama M, et al. Deep whole-genome sequencing reveals recent selection signatures linked to evolution and disease risk of Japanese. Nat Commun. 2018;9:1631.

4. Wu D, Dou J, Chai X, Bellis C, Wilm A, Shih CC, et al. Large-Scale Whole-Genome Sequencing of Three Diverse Asian Populations in Singapore. Cell. 2019;179:736–49.e15.

5. Cao Y, Li L, Xu M, Feng Z, Sun X, Lu J, et al. The ChinaMAP analytics of deep whole genome sequences in 10,588 individuals. Cell Res. 2020;30:717–31.

6. Wei C-Y, Yang J-H, Yeh E-C, Tsai M-F, Kao H-J, Lo C-Z, et al. Genetic profiles of 103,106 individuals in the Taiwan Biobank provide insights into the health and history of Han Chinese. NPJ Genom Med. 2021;6:10.

7. Yang Z, Cui Q, Zhou W, Qiu L, Han B. Comparison of gene mutation spectrum of thalassemia in different regions of China and Southeast Asia. Mol Genet Genomic Med. 2019;7:e680.

8. Onore ME, Torella A, Musacchia F, D’Ambrosio P, Zanobio M, Del Vecchio Blanco F, et al. Linked-Read Whole Genome Sequencing Solves a Double DMD Gene Rearrangement. Genes [Internet]. 2021;12. Available from: http://dx.doi.org/10.3390/genes12020133

9. Shang X, Peng Z, Ye Y, Asan, Zhang X, Chen Y, et al. Rapid Targeted Next-Generation Sequencing Platform for Molecular Screening and Clinical Genotyping in Subjects with Hemoglobinopathies. EBioMedicine. 2017;23:150–9.

10. Chen X, Sanchis-Juan A, French CE, Connell AJ, Delon I, Kingsbury Z, et al. Spinal muscular atrophy diagnosis and carrier screening from genome sequencing data. Genet Med. 2020;22:945–53.

11. Juang J-MJ, Lu T-P, Su M-W, Lin C-W, Yang J-H, Chu H-W, et al. Rare variants discovery by extensive whole-genome sequencing of the Han Chinese population in Taiwan: Applications to cardiovascular medicine. J Advert Res. 2021;30:147–58.

12. Edwards JG, Feldman G, Goldberg J, Gregg AR, Norton ME, Rose NC, et al. Expanded carrier screening in reproductive medicine—points to consider: a joint statement of the American College of Medical Genetics and Genomics, American College of Obstetricians and Gynecologists, National Society of Genetic Counselors, Perinatal Quality Foundation, and Society for Maternal-Fetal Medicine. Obstetrics & Gynecology. LWW; 2015;125:653–62.

13. Miller DT, Lee K, Chung WK, Gordon AS, Herman GE, Klein TE, et al. ACMG SF v3. 0 list for reporting of secondary findings in clinical exome and genome sequencing: a policy statement of the American College of Medical Genetics and Genomics (ACMG). Genet Med. Nature Publishing Group; 2021;1–10.

14. Tang CS-M, Dattani S, So M-T, Cherny SS, Tam PKH, Sham PC, et al. Actionable secondary findings from whole-genome sequencing of 954 East Asians. Hum Genet. 2018;137:31–7.

15. Kuo C-W, Hwu W-L, Chien Y-H, Hsu C, Hung M-Z, Lin I-L, et al. Frequency and spectrum of actionable pathogenic secondary findings in Taiwanese exomes. Mol Genet Genomic Med. 2020;8:e1455.

16. eMERGE Clinical Annotation Working Group. Frequency of genomic secondary findings among 21,915 eMERGE network participants. Genet Med. 2020;22:1470–7.

17. Freed D, Aldana R, Weber JA, Edwards JS. The Sentieon Genomics Tools-A fast and accurate solution to variant calling from next-generation sequence data. BioRxiv [Internet]. biorxiv.org; 2017; Available from: https://www.biorxiv.org/content/10.1101/115717v2.abstract

18. Van der Auwera GA, Carneiro MO, Hartl C, Poplin R, Del Angel G, Levy-Moonshine A, et al. From FastQ data to high confidence variant calls: the Genome Analysis Toolkit best practices pipeline. Curr Protoc Bioinformatics. Wiley; 2013;43:11.10.1–11.10.33.

19. Li H. Aligning sequence reads, clone sequences and assembly contigs with BWA-MEM [Internet]. arXiv [q-bio.GN]. 2013. Available from: http://arxiv.org/abs/1303.3997

20. Danecek P, Bonfield JK, Liddle J, Marshall J, Ohan V, Pollard MO, et al. Twelve years of SAMtools and BCFtools. Gigascience [Internet]. 2021;10. Available from: http://dx.doi.org/10.1093/gigascience/giab008

21. Li H. A statistical framework for SNP calling, mutation discovery, association mapping and population genetical parameter estimation from sequencing data. Bioinformatics. 2011;27:2987–93.

22. Wang K, Li M, Hakonarson H. ANNOVAR: functional annotation of genetic variants from high-throughput sequencing data. Nucleic Acids Res. 2010;38:e164.

23. Chen X, Schulz-Trieglaff O, Shaw R, Barnes B, Schlesinger F, Källberg M, et al. Manta: rapid detection of structural variants and indels for germline and cancer sequencing applications. Bioinformatics. 2016;32:1220–2.

24. Geoffroy V, Herenger Y, Kress A, Stoetzel C, Piton A, Dollfus H, et al. AnnotSV: an integrated tool for structural variations annotation. Bioinformatics. 2018;34:3572–4.

25. Chen P-L, Shih S-R, Wang P-W, Lin Y-C, Chu C-C, Lin J-H, et al. Genetic determinants of antithyroid drug-induced agranulocytosis by human leukocyte antigen genotyping and genome-wide association study. Nat Commun. 2015;6:7633.

26. Numanagic I, Malikic S, Ford M, Qin X, Toji L, Radovich M, et al. Allelic decomposition and exact genotyping of highly polymorphic and structurally variant genes. Nat Commun. 2018;9:828.

27. Lee SB, Wheeler MM, Patterson K, McGee S, Dalton R, Woodahl EL, et al. Stargazer: a software tool for calling star alleles from next-generation sequencing data using CYP2D6 as a model. Genet Med. 2019;21:361–72.

28. Landrum MJ, Lee JM, Benson M, Brown GR, Chao C, Chitipiralla S, et al. ClinVar: improving access to variant interpretations and supporting evidence. Nucleic Acids Res. 2018;46:D1062–7.

29. Karczewski KJ, Francioli LC, Tiao G, Cummings BB, Alfoldi J, Wang Q, et al. The mutational constraint spectrum quantified from variation in 141,456 humans. Nature. 2020;581:434–43.

30. Nagasaki M, Yasuda J, Katsuoka F, Nariai N, Kojima K, Kawai Y, et al. Rare variant discovery by deep whole-genome sequencing of 1,070 Japanese individuals. Nat Commun. 2015;6:8018.

31. Westemeyer M, Saucier J, Wallace J, Prins SA, Shetty A, Malhotra M, et al. Clinical experience with carrier screening in a general population: support for a comprehensive pan-ethnic approach. Genet Med. 2020;22:1320–8.

32. Zhao S, Wang W, Wang Y, Han R, Fan C, Ni P, et al. NGS-based spinal muscular atrophy carrier screening of 10,585 diverse couples in China: a pan-ethnic study. Eur J Hum Genet. 2021;29:194–204.

33. Feng Y, Ge X, Meng L, Scull J, Li J, Tian X, et al. The next generation of population-based spinal muscular atrophy carrier screening: comprehensive pan-ethnic SMN1 copy-number and sequence variant analysis by massively parallel sequencing. Genet Med. 2017;19:936–44.

34. Caillou B, Dupuy C, Lacroix L, Nocera M, Talbot M, Ohayon R, et al. Expression of reduced nicotinamide adenine dinucleotide phosphate oxidase (ThoX, LNOX, Duox) genes and proteins in human thyroid tissues. J Clin Endocrinol Metab. 2001;86:3351–8.

35. De Deken X, Wang D, Many MC, Costagliola S, Libert F, Vassart G, et al. Cloning of two human thyroid cDNAs encoding new members of the NADPH oxidase family. J Biol Chem. 2000;275:23227–33.

36. Moreno JC, Bikker H, Kempers MJE, van Trotsenburg ASP, Baas F, de Vijlder JJM, et al. Inactivating mutations in the gene for thyroid oxidase 2 (THOX2) and congenital hypothyroidism. N Engl J Med. 2002;347:95–102.

37. Vigone MC, Fugazzola L, Zamproni I, Passoni A, Di Candia S, Chiumello G, et al. Persistent mild hypothyroidism associated with novel sequence variants of the DUOX2 gene in two siblings. Hum Mutat. 2005;26:395.

38. Ueyama H, Kuwayama S, Imai H, Tanabe S, Oda S, Nishida Y, et al. Novel missense mutations in red/green opsin genes in congenital color-vision deficiencies. Biochem Biophys Res Commun. 2002;294:205–9.

39. Lu YB, Kobayashi K, Ushikai M, Tabata A, Iijima M, Li MX, et al. Frequency and distribution in East Asia of 12 mutations identified in the SLC25A13 gene of Japanese patients with citrin deficiency. J Hum Genet. 2005;50:338–46.

40. Kobayashi K, Bang Lu Y, Xian Li M, Nishi I, Hsiao K-J, Choeh K, et al. Screening of nine SLC25A13 mutations: their frequency in patients with citrin deficiency and high carrier rates in Asian populations. Mol Genet Metab. 2003;80:356–9.

41. Song Y-Z, Deng M, Chen F-P, Wen F, Guo L, Cao S-L, et al. Genotypic and phenotypic features of citrin deficiency: five-year experience in a Chinese pediatric center. Int J Mol Med. 2011;28:33–40.

42. Kabashima K, Sakabe J-I, Yamada Y, Tokura Y. “Nagashima-Type” Keratosis as a Novel Entity in the Palmoplantar Keratoderma Category. Arch Dermatol. American Medical Association; 2008;144:375–9.

43. Kubo A, Shiohama A, Sasaki T, Nakabayashi K, Kawasaki H, Atsugi T, et al. Mutations in SERPINB7, encoding a member of the serine protease inhibitor superfamily, cause Nagashima-type palmoplantar keratosis. Am J Hum Genet. 2013;93:945–56.

44. Cottin V, Thibout Y, Bey-Omar F, Durieu I, Laoust L, Morel Y, et al. Late CF caused by homozygous IVS8-5T CFTR polymorphism. Thorax. 2005;60:974–5.

45. Wu C-C, Alper OM, Lu J-F, Wang S-P, Guo L, Chiang H-S, et al. Mutation spectrum of the CFTR gene in Taiwanese patients with congenital bilateral absence of the vas deferens. Hum Reprod. 2005;20:2470–5.

46. Huang H, Xu L, Chen M, Lin N, Xue H, Chen L, et al. Molecular characterization of thalassemia and hemoglobinopathy in Southeastern China. Sci Rep. 2019;9:3493.

47. He J, Song W, Yang J, Lu S, Yuan Y, Guo J, et al. Next-generation sequencing improves thalassemia carrier screening among premarital adults in a high prevalence population: the Dai nationality, China. Genet Med. 2017;19:1022–31.

48. Zhang J, Wang Y, Ma D, Sun Y, Li Y, Yang P, et al. Carrier Screening and Prenatal Diagnosis for Spinal Muscular Atrophy in 13,069 Chinese Pregnant Women [Internet]. The Journal of Molecular Diagnostics. 2020. p. 817–22. Available from: http://dx.doi.org/10.1016/j.jmoldx.2020.03.001

49. Dejsuphong D, Taweewongsounton A, Khemthong P, Chitphuk S, Stitchantrakul W, Sritara P, et al. Carrier frequency of spinal muscular atrophy in Thailand [Internet]. Neurological Sciences. 2019. p. 1729–32. Available from: http://dx.doi.org/10.1007/s10072-019-03885-5

50. Park JE, Yun SA, Roh EY, Yoon JH, Shin S, Ki C-S. Carrier Frequency of Spinal Muscular Atrophy in a Large-scale Korean Population [Internet]. Annals of Laboratory Medicine. 2020. p. 326–30. Available from: http://dx.doi.org/10.3343/alm.2020.40.4.326

